# The role of childrens’ vaccination for COVID-19 - Pareto-optimal allocations of vaccines

**DOI:** 10.1101/2021.04.26.21256101

**Authors:** Nir Gavish, Guy Katriel

## Abstract

The ultimate goal of COVID-19 vaccination campaigns is to enable the return of societies and economies to a state of normality. While vaccines have been approved for children of age 12 and older, there is an ongoing debate as to whether children should be vaccinated and at what priority, with very different policies being adopted in different countries. In this work, we use mathematical modeling and optimization to study the effect of vaccinating children on the epidemic spread. We consider Pareto-optimal allocations according to competing measures of number of infections and mortality, and systematically study the trade-offs among them. When some weight is given to the number of infections, we find that it is optimal to allocate vaccines to adolescents in age group 10-19, even when they are assumed to be less susceptible than adults. Additionally, we find that in a broad range of scenarios, optimal allocations of vaccines do not include vaccination of age-group 0-9.

**Author summary:** One of the acute questions public health experts and policymakers currently confront is whether children of age 12 and older, and eventually perhaps younger children, should be vaccinated against COVID-19, and at what priority. Different countries have adopted diverse policies on this issue, while others remain undecided. One of the key considerations in this debate is the impact of children’s vaccination on the epidemic spread. In this work, we use mathematical and computational methods to study this question in a systematic, quantitative way. We compute optimal vaccination allocations, under different criteria for optimality, both including and not including children. To explore tradeoffs among different goals, such as reducing number of infections and reducing mortality, we use the idea of Pareto optimization, which is novel in this field. Our results show that, under a wide range of conditions, optimal vaccine allocations include vaccination of age group 10-19, while the vaccination of age group 0-9 is of lower priority than the vaccination of other age group.

## 1 Introduction

The availability of effective vaccines against SARS-CoV-19 is widely seen as a game-changer that will enable an eventual return to normal life in those countries in which it becomes available, following the devastating COVID-19 pandemic. Presently vaccines have been approved for children of age 12 and older, but the question of whether children should receive COVID vaccines remain under debate [1]. At the time of submission, the US and Canada have included all eligible children in their vaccination campaign, while European countries are almost evenly split on whether to administer coronavirus vaccinations to adolescents, with several European countries remaining undecided. With ongoing vaccine trials for children under 12, the question of vaccinating younger children will likely come to the fore in coming months [2, 3].

A key question under consideration in this debate is whether and to what extent vaccination of children will enhance the effectiveness of a vaccination campaign at the population level [4–6]. When attempting to assess the relative merits of allocating vaccines to the younger age groups, one must take into account the epidemiological characteristics of COVID-19 with respect to these groups, and these seem to point in opposing directions. On the one hand, children infected with COVID-19 rarely develop severe disease [7, 8]. In addition, it has been estimated that children’s susceptibility to infection by SARS-CoV-19 is significantly lower than that of adults [9–11]. On the other hand, children are a relatively large age group that tends to interact more intensively than other age groups. A large epidemic outbreak among children is likely to spread to older age groups and risk vaccinated and non-vaccinated adults, so that vaccination of children indirectly protects individuals of other age groups [12], who are at greater risk of severe outcomes. These considerations suggest that, in the case of COVID-19, quantifying the *indirect* effect of vaccinating the younger age groups in protecting the older, more vulnerable age-groups is essential for evaluating the benefits of allocating vaccines to children and adolescents. Ethicists have been debating whether vaccinating children is justified, under the appropriate circumstances, even if the primary aim of doing so is to protect the older age groups [13, 14]. However this debate is only relevant if indeed allocating vaccines to children enables achieving better outcomes than allocating them to older age groups.

The goal of this work is to contribute to the discussion on childrens’ vaccination by quantifying the population level impact of childrens’ vaccination. We present a mathematical model to explore the effect of demography, age-based social interaction structure, and vaccine efficacy on the optimal post-vaccination outcomes that can be achieved by suitable allocation of vaccines, in the context of the COVID-19 pandemic. Our study focuses on two questions:

1. How essential is the vaccination of children and youths to achieving herd immunity? Specifically, what are the prospects for achieving herd immunity with the aid of vaccination, assuming vaccine allocation is performed optimally, and what are the age-dependent vaccine allocations which will achieve herd immunity with minimal vaccination coverage?
2. What is the population level impact of vaccination of children in case herd immunity by vaccination cannot be attained? What are the optimal outcomes - according to different possible measures - that can be achieved, and what are the age-dependent vaccine allocations that will achieve them?

To address these questions we compare scenarios in which all age-groups are eligible, by policy, for vaccination, to scenarios in which vaccination is restricted only to those over 10, or only to those over 20. When children *can* be vaccinated, we wish to determine whether it is *optimal* to do so, under conditions of limited vaccine availability. When children *cannot* be vaccinated, we seek to evaluate whether and to what extent the optimal achievable outcomes are degraded relative to the case in which children can be vaccinated.

Several recent model-based studies address the question of optimal vaccine allocations for SARS-Cov-19. Moore et al. [15] studied vaccination strategies for COVID with the aim of minimizing future deaths or quality adjusted life year losses. This study was conducted at an early stage of the pandemic when uncertainty regarding the vaccines was high. Bubar et al. [16] focus on the design of a vaccination campaign as it competes with the spread of infection. Matrajt et al. [17] as well Meehan et al. [18], used an age-stratified model to study the consequences of vaccine effectiveness and population coverage on the optimal vaccine allocation. These works show that when available vaccine coverage is relatively low, mortality-minimizing vaccine allocations prioritize the elderly, while for sufficiently high coverage the mortality-minimizing vaccine allocations are those that prioritize younger populations who are the drivers of the epidemic. Vaccine allocations that are optimized according to other criteria which are correlated with mortality, e.g., ICU peak, give rise to qualitatively similar patterns, but the point of transition between vaccination of the young and vaccination of the elderly varies.

In this work, we investigate outcomes of a vaccination campaign in the medium-term range, after the vaccination effort has ended. Our work is complementary to the above mentioned studies [15–18] in both its focus and methods used. We specifically address the issue of restrictions on the eligibility of children and adolescents, and quantify the consequences of choosing not to vaccinate them. An additional novel aspect of this work is the simultaneous consideration of several objectives of a vaccination campaign, by employing the concept of Pareto-optimality. Vaccination policies are commonly optimized with respect to a single measure such as mortality, number of infections, quality adjusted life year losses or hospitalizations [15–19]. Consequently, vaccination studies present multiple optimal strategies, each optimized with respect to a different measure. However, from a policy maker point of view, it is not clear 1) which optimal allocation should be chosen, 2) what are the trade-offs between the measures when determining an allocation, 3) how robust is the choice of allocation to changes in assumptions. Here, we address these issues, by taking a different approach and considering the problem as one of multi-objective optimization, giving rise to a set of Pareto-optimal vaccine allocations. Given two (or more) measures for the outcome of a vaccination campaign (*e*.*g*. mortality and number of infections), an allocation is called *Pareto optimal* (with respect to these measure) if there is no other allocation which produces an outcome which is better with respect to all measures. Assuming that the measures employed faithfully characterize our criteria for evaluating outcomes, it would be irrational to chose an allocation which is *not* Pareto-optimal. While there are infinitely many Pareto-optimal allocations, and corresponding outcomes, so that the choice of a specific one among them requires a weighing of the different outcome measures, the Pareto-optimality approach allows to systematically evaluate the trade-offs between competing measures such as mortality and number of infections. In addition, by comparing the possible outcomes of Pareto-optimal allocations (the *Pareto front*) for the scenarios in which all age-groups can be vaccinated with those obtained when young age groups are not vaccinated, we obtain a global view of the extent of the effect that limiting eligibility for vaccination has on the outcomes. More generally, such comparisons among Pareto fronts provide a a way to visualize the impact of various changes in assumptions.

## 2 Methods

This section presents the mathematical model that was used as a basis for this study, and the analytical and numerical methods that were used in order to explore questions related to optimal vaccination using the model.

### 2.1 Age-structured SIR model with vaccination

The computations in this work rely on an age-stratified SIR model [16, 20, 21]. It should be mentioned that since our results concern only the herd-immunity threshold and final sizes, and since these quantities do not depend on the generation-time distribution [21], the results derived are identical to those that would be obtained from a more elaborate SEIR or a more general age-of-infection model.

The population is divided into *n* age groups. The dynamic variables are *S*_*j*_, *I*_*j*_, *R*_*j*_ and *V*_*j*_, the numbers of susceptible, infected, recovered and vaccinated individuals in age-group *j* (1 ≤ *j* ≤ *n*).

Parameters of the model are:

- *N*_*j*_ (1 ≤ *j* ≤ *n*) is the size of age group *j*.
- *C*_*jk*_ denotes the mean number of contacts of a single member of age group *j* with members of group *k* per unit time. We denote by *C* the *n × n* matrix with elements 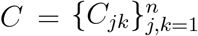
- *β*_*j*_ is the probability of infection upon contact for members of group *j*, allowing for varying susceptibility to infection in different age groups.
- *γ* denotes the recovery rate, so that 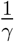 is the mean duration of infectivity.
- *p*_*j*_ *∈* [0, 1] is the fraction of group *j* which is vaccinated.
- *ν* is the fraction of those vaccinated for which protective immunity is generated.
- 1 − *ε* is the vaccine efficacy against infection, so that *ε* is the factor by which probability of infection upon contact is reduced for those vaccinated.

The case *ε >* 0, *ν* = 1 is known as ‘leaky vaccine’, and the case *ε* = 0, *ν <* 1 is known as an ‘all-or-none’ vaccine.

The dynamics is then described by the differential equations

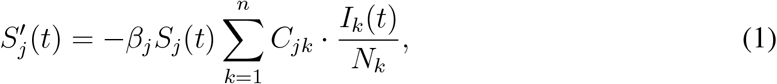

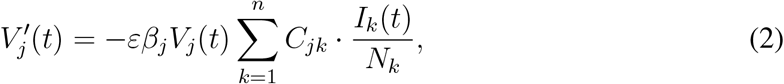

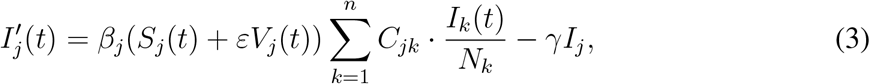

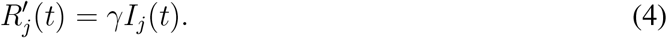

We assume that the initial numbers of infected *I*_*j*_ (0) and of recovered *R*_*j*_ (0) are given. Since a proportion *p*_*j*_ of age group *j* is vaccinated, and a fraction *ν* of these will generate immunity, we have

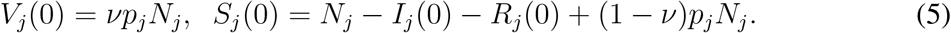

To calculate the basic reproductive number, *R*_0_, we use the next-generation matrix

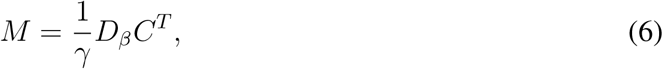

where *D*_*β*_ is the diagonal matrix with diagonal entries *β*_*i*_ and *C* is the country-specific contact matrix. The basic reproductive number *R*_0_ is equal to *ρ*(*M*), the spectral radius of *M* [20, 21].

### 2.2 Parameter values

Here we describe the parameter values used in the computations which were carried out.

- **Age Demographics** in all simulations were taken from the UN World Population Prospects 2019 for each country [22], using *n* = 9 age groups, of sizes *N*_*j*_ (1 ≤ *j* ≤ 9) corresponding to 10-year increments, with the last group comprising those of age 80 and older.
- **Contact matrices** 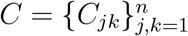. We used contact matrices from [23]. Age bins in each case were originally provided in 5-year increment, where the last age bin corresponds to ages 75 and older. We follow the procedure as in [16] to adapt the matrices into 10-year increments.
- **Susceptibility parameters** *β*_*i*_ : The examples presented in this study assume the age dependent susceptibility profile for SARS-CoV-19 from [9]:

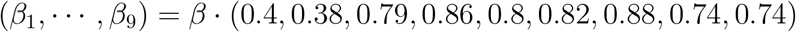

in which the relative susceptibility of age group 0-19 is roughly half of older age groups. The parameter *β* is adjusted to obtain different values of *R*_0_.
- **Vaccine efficacy** Unless otherwise specified, in what follows, we assume that the susceptibility of vaccinated individuals to infection is reduced by 80% (so *ε* = 0.2), and the risk of a vaccinated infectee to develop severe disease is 25% that of a non-vaccinated infectee. By construction, this combination of parameters gives rise to an overall reduction of 95% in the risk of a vaccinated individual to develop severe disease as estimated in controlled studies [24] and analysis of real-world data [25]. The fraction *ν* is the fraction of those vaccinated for which protective immunity is generated is taken to be *ν* = 1.
- **Age-dependent infection fatality ratio (IFR)** We assume the age dependent IFR profile [16, 26],

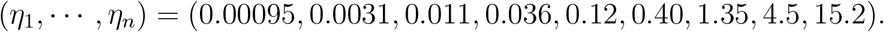
- **Initial values**: The initial values for the differential equations, which are used in the final-size formulas, are *R*_*i*_ (0) = 0 for 1 ≤ *i* ≤ *n*, unless otherwise stated, that is we assume no recovered individuals. We also take *I*_*i*_ (0) = 0 is the final size formula - corresponding to a very small fraction of the population initially infected.

We note that, as far as the computations performed here are concerned, the value of parameter *γ* (recovery rate) in the model has no effect, since in the expressions for the reproductive number, as well as in the final size equations, 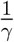 is multiplied by the parameter *β*, which is adjusted to achieve the desired value of *R*_0_. Therefore we do not need to fix a value for the parameter *γ*.

### 2.3 Computation of vaccine supply threshold

The post-vaccination effective reproduction number *R*_*v*_ is the spectral radius *ρ*(*M*_*v*_) of the nextgeneration matrix following vaccination

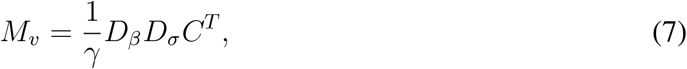

where *D*_*σ*_ is a diagonal matrix with diagonal entries

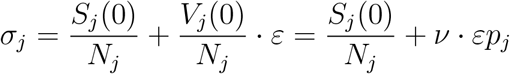

The matrix *M*_*v*_ depends on the vaccination fractions *p*_*j*_ in each age group, and to stress this we will denote it by *M*_*v*_ (*p*_1_, *…, p*_*n*_).

The vaccine supply threshold is the *minimal* vaccine coverage required for achieving herd immunity, that is attaining *R*_*v*_ = 1 [27, 28]. To compute this quantity we define, for each level 0 ≤ *p* ≤ 1 of total vaccine coverage, the minimal reproductive number attainable using allocations with total coverage *p*, that is

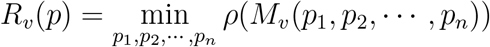

subject to the constraints

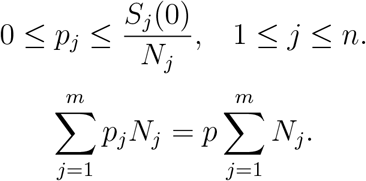

The minimal vaccination coverage required to achieve herd immunity is obtained by solving the equation *R*_*v*_ (*p*) = 1, and the corresponding minimizer (*p*_1_, *p*_2_, *…, p*_*n*_) gives the optimal vaccine allocation for achieving herd immunity.

We conduct these computations using Matlabs’ `fmincon` nonlinear programming solver with a sequential quadratic programming algorithm, where the vaccine coverage *p* is gradually increased and the initial guess used for coverage *p* + *δp* is adapted from the solution of the optimization problem for vaccine coverage *p*. The choice of the sequential quadratic programming algorithm is more suitable than the default interior point algorithm used by `fmincon`, since the initial guess provided typically lies on the boundary of the feasible set rather than in its interior. We should note that in general the optimization problem that we solve here is a non-convex one [27, 28], so we do not have theoretical guarantees that there will not exist local minimas, at which the optimization algorithm could get stuck without finding the global minimum. To reduce the probability of convergence to a local minimum, we have randomized the initial point provided to the algorithm and verified that it converges to the same minimum, so that we are reasonably confident that we have found the global minima.

### 2.4 Final size formula

The final-size formula [20,21] yields the overall number of infections in each age group in terms of the model parameters, allowing us to compute the epidemic’s outcome without the need to numerically solve the differential equations.

To derive the final-size formula for the current model, it is convenient to reformulate the system (1)-(4) in terms of proportions

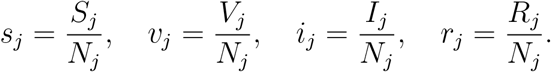

Obtaining

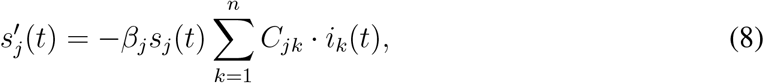

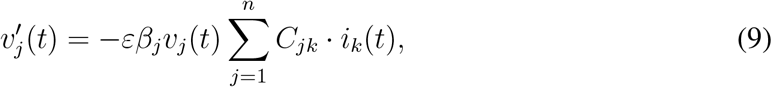

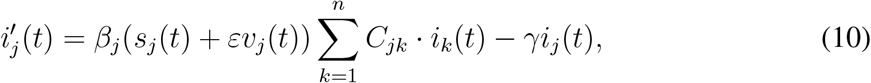

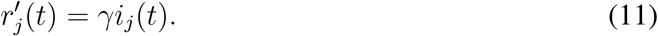

Assuming that the initial fractions of infected *i*_*j*_ (0) and of recovered *r*_*j*_ (0) are given, (5) translates into

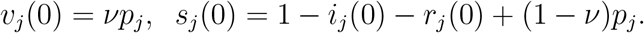

From (8),(9) we have

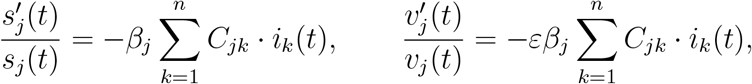

which upon integration yields

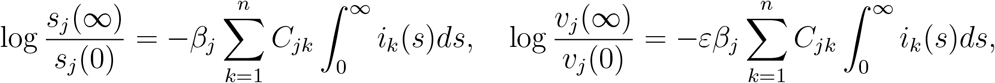

or

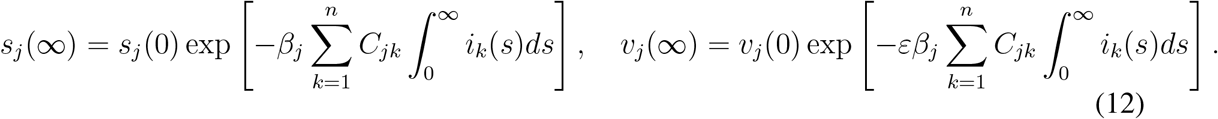

Note that, assuming *s*_*i*_ (0) ≠ 0, (12) implies the relation

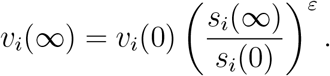

Summing (8)-(10), we have

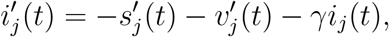

which, upon integration, gives

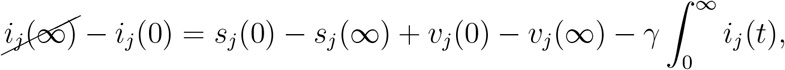

or

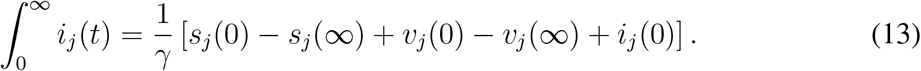

Since *s*_*j*_ (*t*) + *v*_*j*_ (*t*) + *i*_*j*_ (*t*) + *r*_*j*_ (*t*) = 1 for all *t*, and *i*_*j*_ (∞) = 0, we have

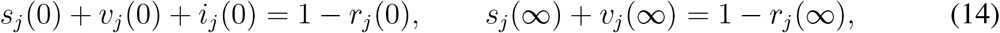

and can write (13) as

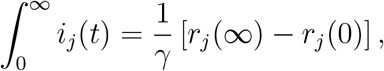

so that (12) yields

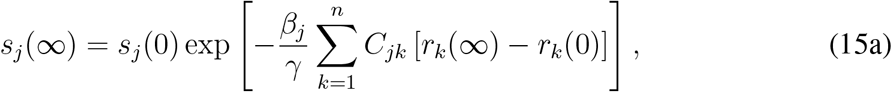

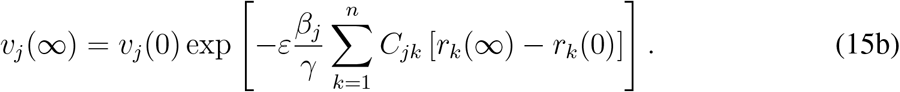

Combining (14) and (15) yields

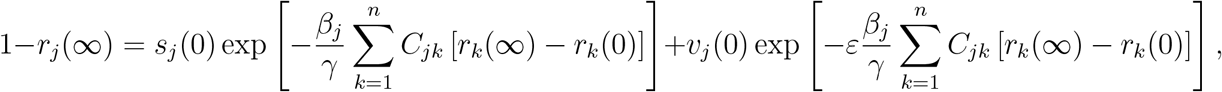

or, defining *z*_*j*_ = *r*_*j*_ (∞)−*r*_*j*_ (0) to be fraction of group *j* infected throughout the post-vaccination period:

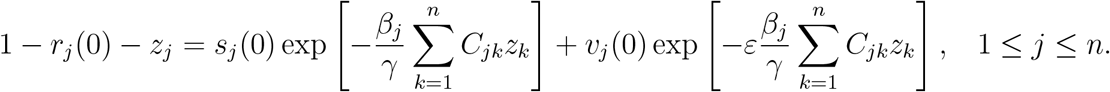

Solving this system of equation numerically yields the fractions *z*_*j*_.

As an example, we compute the final size of an epidemic spreading after 55% of the population of the USA is vaccinated with no age prioritization, i.e., 55% of each age group is vaccinated. Considering a post-COVID basic reproduction number of *R*_0_ = 3, we observe that by the end of the epidemic 25.8% of the population above the age of 80 will be infected without the protection of a vaccine, see Figure 1A. When vaccines are homogeneously allocated only to adults (ages 20 and over), the portion of the population above the age of 80 that is infected without the protection of a vaccine drops to 15.7%, but 78.9% of children in the age group of 10-19 are infected, see Figure 1B. The above examples assume that the entire population is either susceptible or vaccinated at the end of the vaccination campaign. In order to model in a more realistic manner we allow for preexisting immunity due to recovery, as well as for the prevalence of active cases, see Figure 1C.

**Figure 1:**
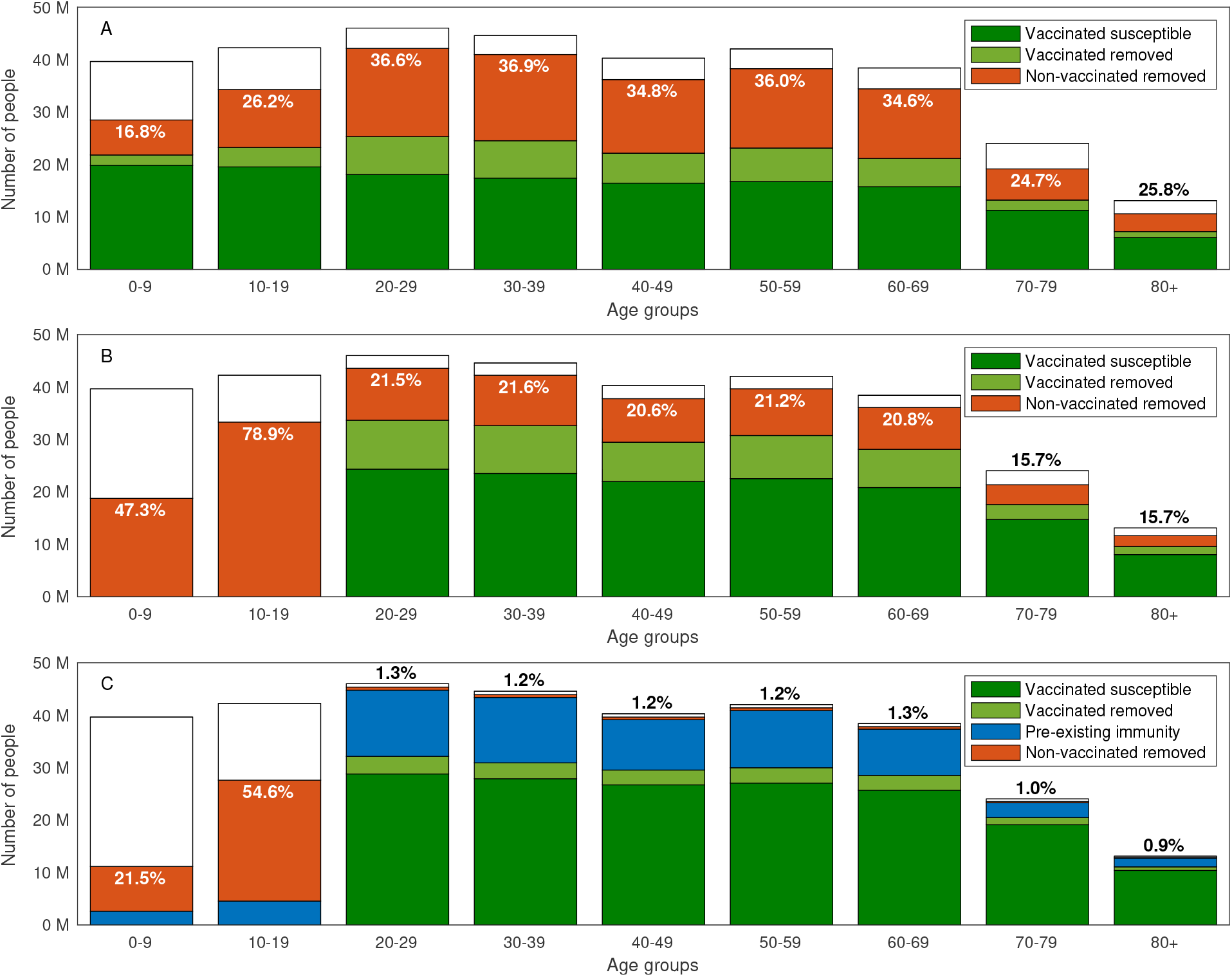
Final size of epidemic. Final size of an epidemic spreading with basic reproduction number of *R*_0_ = 3 after 55% of the population of the USA is vaccinated with no age prioritization. Removed population refers to those recovered or dead. The computation considers a vaccination campaign in which A: Vaccines are allocation to all ages. B: Vaccine allocation is limited to ages 20 and above. C: Vaccine allocation is limited to ages 20 and above, 20% of the population has pre-existing immunity recovered from COVID-19, and the prevalence of active cases is 0.5% of the population. The text in all graphs corresponds to the percent of non-vaccinated removed individuals in each age group.

### 2.5 Computation of optimal vaccine allocations

For a given vaccine allocation 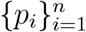 where *p*_*i*_ is the fraction of age group *i* which is vaccinated, we use the final size formula to compute the outcomes in terms of the fraction of each age group infected *z*_*j*_ (1 ≤ *j* ≤ *n*). The function *f* (*p*_1_, *p*_2_, *…, p*_*n*_) to be minimized in the case that the aim is to minimize the number of infections is

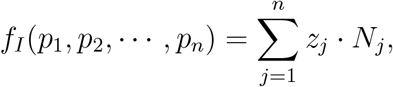

while if the aim is minimizing mortality we take

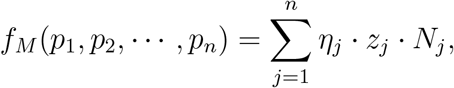

where *η*_*j*_ (1 ≤ *j* ≤ *n*) are the infection fatality rates (IFR) in each age group.

Given the total fraction *p* of the population to be vaccinated, we consider the following optimization problem (with *f* = *f*_*I*_, *f* = *f*_*M*_ or a convex combination of these functions):

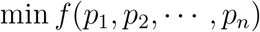

subject to

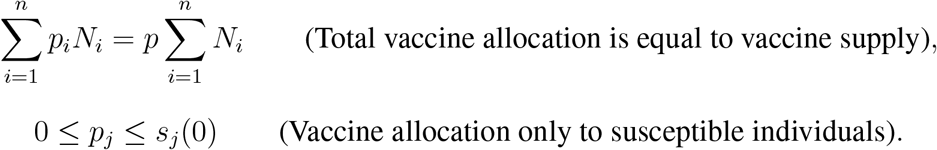

This optimization problem is solved using Matlabs’ `fmincon` nonlinear programming solver.

The inequality constraints can readily be modified so that vaccine allocation also does not exceed a given portion *α*_*j*_ of age group *j*,

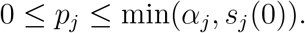

This modification enables to account for age groups for which vaccination is not approved (by setting *α*_*j*_ = 0, as well as for vaccine hesitancy, logistical difficulties in reaching the entire population of an age group, or a portion of the population who cannot be vaccinated due to medical conditions or allergies.

### 2.6 The Pareto front and its computation

The Pareto front is a tool which allows us to examine the trade-offs among competing measures for the effectiveness of a vaccination campaign - in our case the trade-off between minimizing the number of infections (attack rate) and mortality. For a given vaccination coverage, an outcome (*Z, M*) (attack rate and mortality) is called *feasible* if it can be achieved by a suitable allocation of vaccines satisfying the coverage constraint. An outcome is called *Pareto optimal* if it is feasible, and if there do no exist feasible outcomes (*Z*′, *M* ′) which improve upon it both in terms of attack rate and in terms of mortality (*Z*′ *< Z, M* ′ *< M*). The set of Pareto-optimal outcomes is called the *Pareto front*. We aim to compute the Pareto front and display it graphically in the plane of outcomes (*Z, M*).

Each point on the Pareto front corresponds to the mortality minimizing vaccine allocation with a given number of infections. To compute the Pareto front, we first compute its endpoints - namely, we compute the vaccine allocation minimizing attack rate and the vaccine allocation minimizing mortality, with corresponding outcomes (*Z*_0_, *M*_0_) and (*Z*_*L*_, *M*_*L*_), respectively. We conduct these computations using Matlabs’ `fmincon` nonlinear programming solver. To avoid convergence of the optimization algorithm to a local minimum, we run the solver with a set of random initial guesses.

The computation of the end points of the Pareto front determines the range for the attack rate along the Pareto front, and allows to determine a grid of *L* points

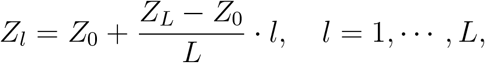

along which the Pareto front is sampled. The optimal allocations along the Pareto front are computed sequentially from one end of the Pareto front to the other by solving the constrained optimization problem of finding the mortality minimizing vaccine allocation with a given number *Z*_*l*_ of infections for *l* = 1, 2, *…, L* − 1:

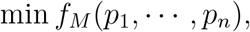

subject to

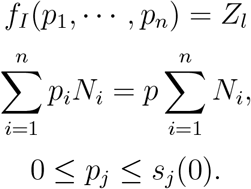

The initial guesses used for each optimization problem at stage *l* is a set of random allocations around the optimal allocation found for the point *l* − 1.

In some cases, we have observed that the direction of sweep from one end to the other affects the results obtained. To eliminate this factor, we sweep in the opposite direction and if needed update the optimal allocation computed. Namely, we recompute the Pareto front at points *Z*_*l*_ for *l* = *L* − 1, *L* − 2, *…*, 1 where the initial guess for each optimization problem is the optimal allocation found for the point *l* + 1.

## 3 Results

In what follows, we consider scenarios of a partial return to normality, to a basic reproduction number of *R*_0_, after vaccination efforts are completed. We illustrate our results using parameters corresponding to the USA demography and contact structure, and later consider other how the different demographic structure of other countries affects the results. Note that *R*_0_ denotes the reproductive number in the absence of vaccination and preexisting immunity due to recovery.

### 3.1 Vaccination coverage required for herd immunity

Achieving herd immunity requires vaccination of a sufficiently large sub-population, and the required coverage can be minimized by allocating the vaccines to different age groups in an optimal manner. Here, for each value of *R*_0_, we compute the minimal vaccine coverage *V*_threshold_ necessary to reach herd immunity, and the corresponding vaccine allocation among the eligible age groups that achieves this goal. See Section 2.3 for details on these computations..

We first examine the case in which the entire population is eligible for vaccination. The results vary with the reproductive number *R*_0_. For example, when *R*_0_ is less than ≈ 1.1, the required vaccine supply is rather small and is allocated solely to age group 30-39, see Figure 2B. Then, as *R*_0_ increases, required vaccine supply *V*_threshold_ gradually increases and its allocation is extended to additional age groups. The optimal vaccine allocations are not necessarily allocations to those who do the most transmitting. Indeed, as *R*_0_ increases, additional age groups are typically added to the allocation before the coverage of the age groups already present in the allocation has reached 100%. For *R*_0_ = 3, herd immunity can be achieved by vaccinating roughly 65% of the population in an optimal way, see Figure 2A. In comparison, if vaccines are allocated *pro rata* (in proportion to the size of age groups), achieving herd immunity requires vaccination of 85% of the population, taking into account 80% vaccine efficacy against infection.

**Figure 2:**
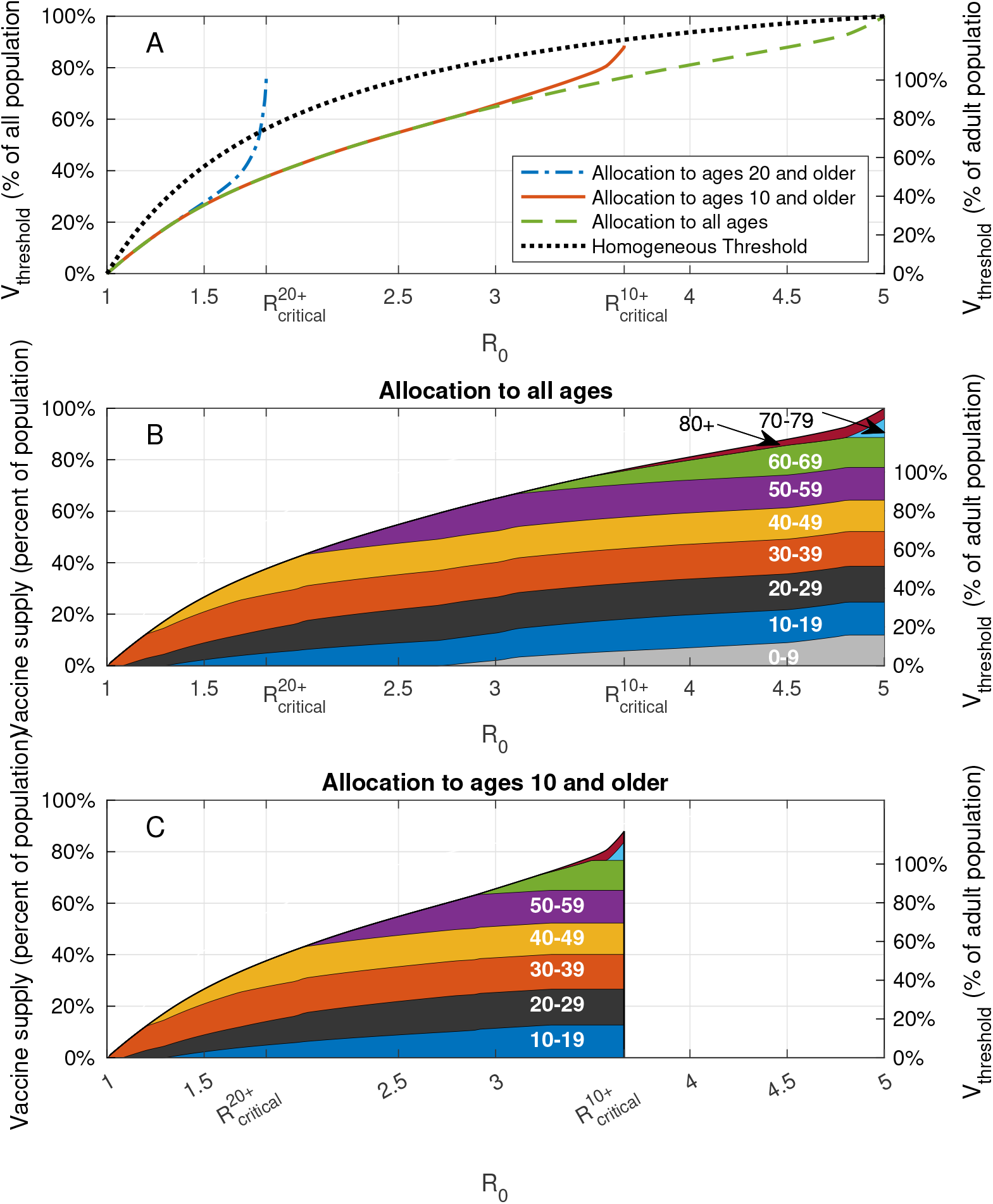
Vaccination coverage required for herd immunity. A: Vaccine coverage *V*_threshold_ required to achieve herd immunity threshold as a function of the reproduction number *R*_0_ for the USA demography and contact structure. B: Vaccine allocations at which herd immunity is achieved at minimal vaccine coverage and when all the population is eligible for vaccination. C: Same as B, but when the population of age 10 and older is eligible for vaccination.

In order to assess the population level impact of the vaccination of children of age 10 and younger on the prospects for achieving herd immunity, we now repeat the analysis while restricting the allocation of vaccines to ages 10 and older, and compute the corresponding threshold curve 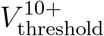 as a function of *R*_0_. We observe that for low values of *R*_0_ the threshold curve 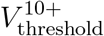 coincides with the threshold curve *V*_threshold_ corresponding to the case in which all ages are vaccine eligible, see Figure 2B in comparison with Figure 2C. The implication is that for values of *R*_0_ *<* 2.75, any allocation achieving herd immunity and including age group 0 − 9 would be *suboptimal* in the sense that its vaccine coverage is larger than the minimum required. For *R*_0_ *>* 2.75, the two curves diverge. This reflects the fact that, if there are no age restrictions, for these values of *R*_0_ all age groups (including children 0 − 9) partake in the optimal allocation (see Figure 2B). Hence, for these values of *R*_0_, any allocation achieving herd immunity and *not* including age group 0 − 9 would be *suboptimal* in that it would require higher level of vaccination overall than the minimal achievable coverage. As *R*_0_ increases further, the threshold curve 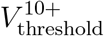 rapidly increases up to its maximal value which corresponds to 100% of the eligible population at 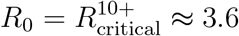. When 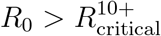, reaching herd immunity becomes *impossible* if children under the age of 10 are not vaccinated.

In case vaccines are not allocated to age group 0-19, we find that herd immunity is achievable only for rather low reproductive numbers, 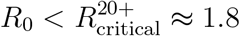. This means that for higher values of *R*_0_, even if *all* adults over 20 were vaccinated, the spread of the infection would be sustained solely by the population under the age of 20. Therefore, for typical values of *R*_0_ of SARS-CoV-19 and its variants, vaccination of age group 10-19 is essential for achieving herd immunity.

The above example, presented in Figure 2, relies on the demographic structure and the contact matrix of the USA. We have also examined the critical reproduction numbers 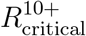 and 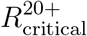 for eight additional countries and found the results to be similar, see Figure 3A. We further observe that the question of whether the infection can be eradicated without vaccinating children is not answered by looking at the percentage of children in the population, as a naive calculation based on a homogeneous-population model would imply. For example, in the case of Zimbabwe, with 53% of its population in age group 0-19, we computed 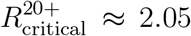, which is higher than 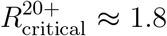 computed for Poland for which the size of age group 0-19 is 20% of the total population. Rather, the key factor is the level of assortativity of mixing within the children sub-population, as reflected in the contact matrix.

**Figure 3:**
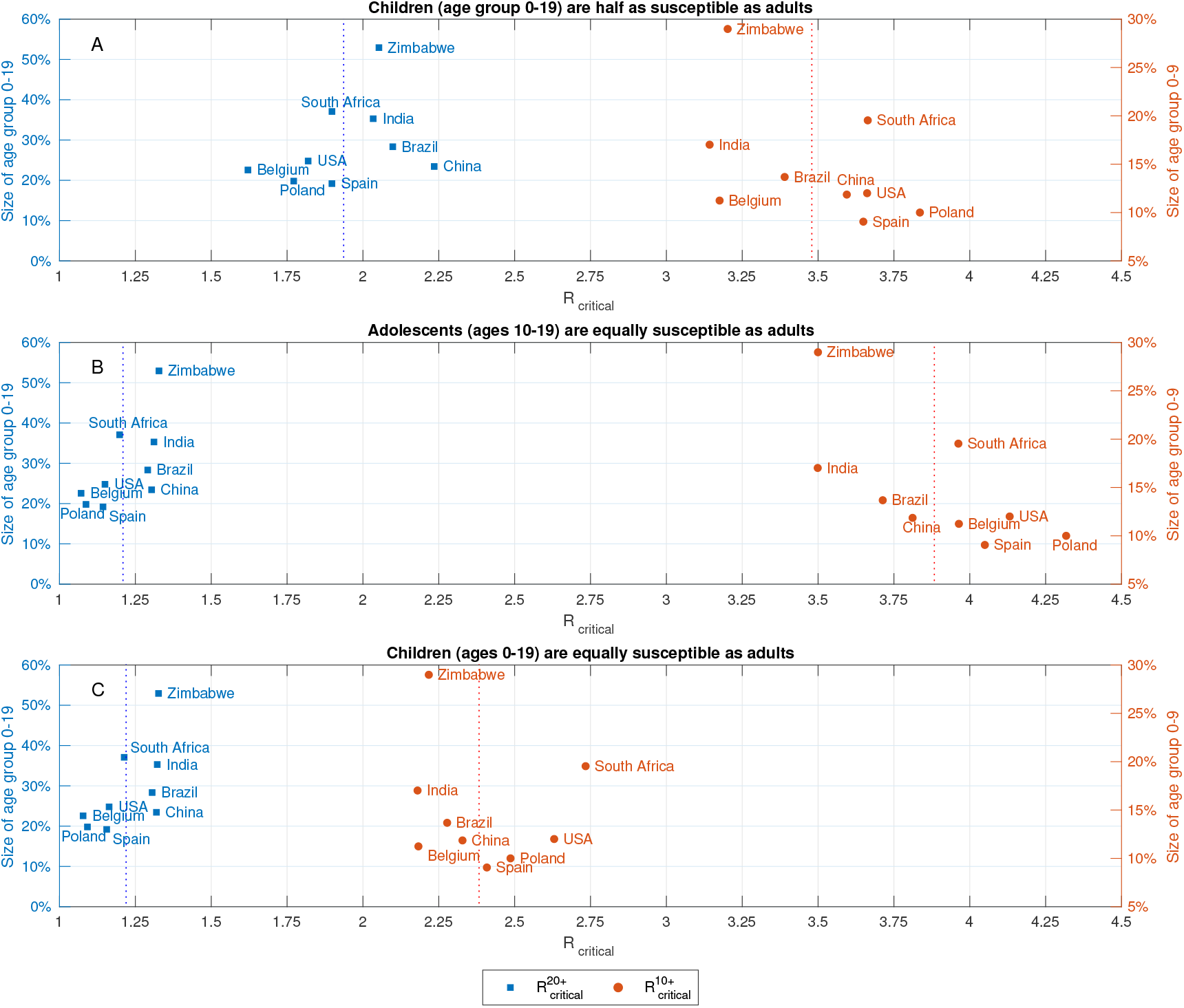
Critical reproduction numbers. Reproduction numbers 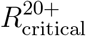 and 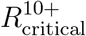 at which herd immunity cannot be achieved without vaccination of age groups 0 − 19 and 0 − 9, respectively. Computed using A: age dependent susceptibility profile of SARS-CoV-19: Ages 0-19 are roughly half as susceptible as adults [9]. B: Same susceptibility profile as in A, but with increased susceptibility in age group 10-19: Ages 10-19 are as susceptible as adults. C: Same susceptibility profile as in A, where ages 10-19 are as susceptible as adults.

### 3.2 Aiming for herd immunity is not a robust strategy

There is considerable uncertainty concerning the value of the reproduction number *R*_0_, as it depends on biological features as well as population-specific attributes such as culture, behavior, living conditions etc. Additionally, *R*_0_ may vary in time, possibly abruptly, both as result of the emergence of viral variants and due to non-pharmaceutical measures in place in the post-vaccination phase. In Section 3.1 above we showed how vaccine coverage increases with *R*_0_ and optimal allocations gradually extend to more age group, see Figure 2. We now examine cases in which reaching herd immunity by vaccination is not feasible, due to limitations on vaccine eligibility or vaccine supply. In such cases, with removal of non-pharmaceutical interventions following the vaccination campaign, herd immunity will eventually be achieved via an epidemic outbreak.

We assess the outcomes of post-vaccination epidemic spread by considering two widely employed measures: attack rate (overall number of infections), and mortality. Attack rate is computed using the final size formula which provides the number of individuals per age group who will be infected, given a the basic reproduction number, see Section 2.4 for details. Expected mortality is then directly computed using an age-dependent infection-fatality ratio. We consider vaccine allocation designed to minimize one of the above measures, see Section 2.5 for details on the computation of such optimal allocations.

Let us first consider allocations minimizing the number of infections, under the assumption that vaccine coverage is 55% of the population, see left part of Figure 4. As long as *R*_0_ is below the herd immunity threshold 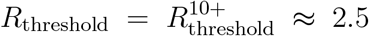, the optimal vaccine allocations for minimizing attack rate coincide with the vaccine allocations that ensure herd immunity below the threshold. For *R*_0_ larger than the herd immunity threshold, herd immunity is not achieved solely by vaccination, but rather also by recovery due to epidemic spread following the vaccination campaign. In this case, we observe that the number of infections and mortality gradually increase with *R*_0_. The structure of the allocations optimized to minimize infections, displayed in the bottom left panel, varies only slightly with *R*_0_. Particularly, as *R*_0_ increases, the allocation prioritizes younger age groups so that at high values of *R*_0_ a small fraction of age group 0-9 is included in the allocation.

**Figure 4:**
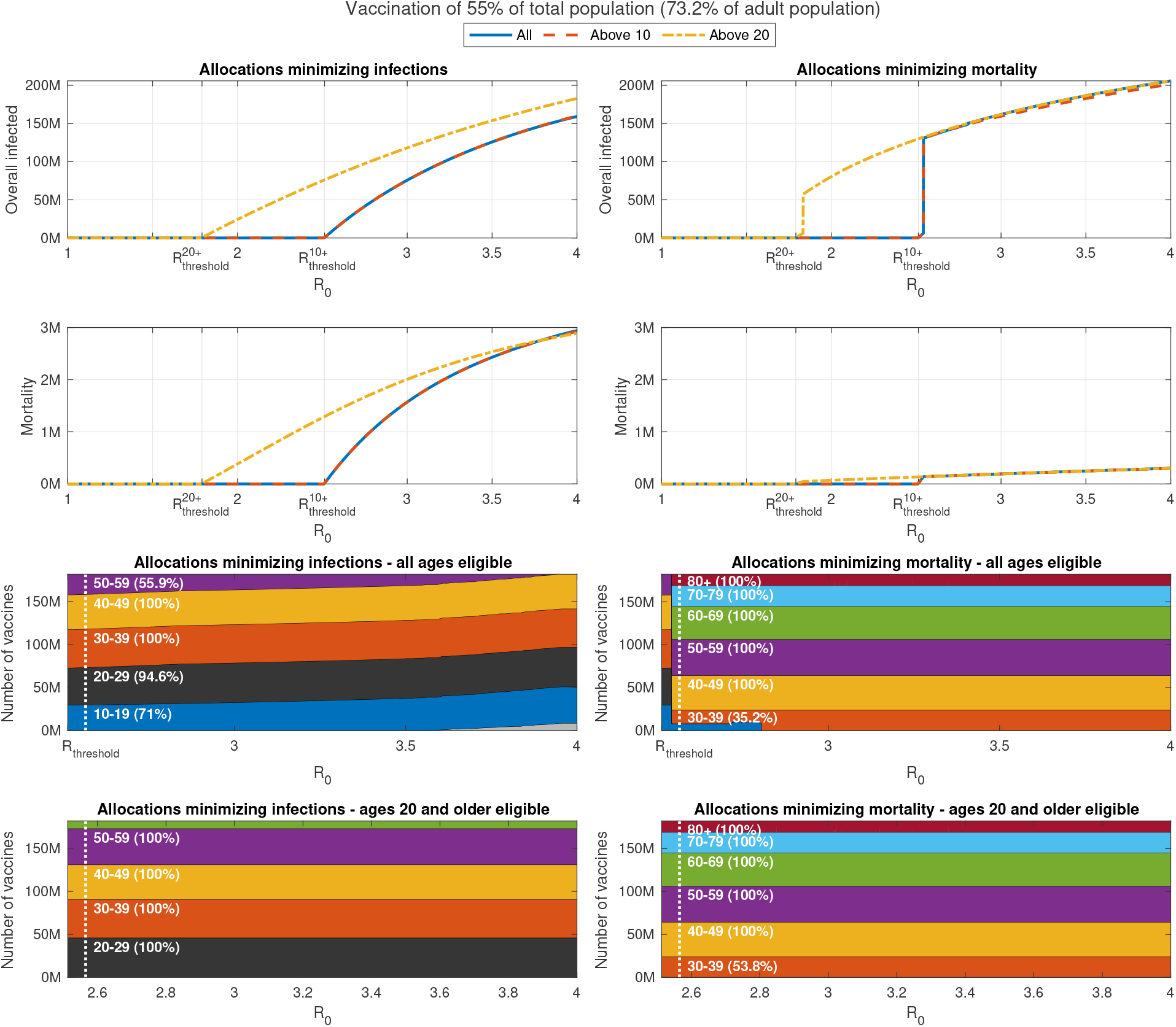
Impact of change in reproduction number. Overall infections of non-vaccinated individuals (top graphs) and overall mortality (centered graphs) as function of the basic reproduction number *R*_0_ after completion of a vaccination campaign for allocations minimizing infections (left panels) and allocations minimizing mortality (right panels). The outcomes are presented for the following cases: All ages are eligible for vaccination (solid), only ages 10 and older are eligible (dashes), and only ages 20 and older are eligible (dash-dots). Bottom panels present allocations when all ages are eligible.

In contrast, for the case of vaccine allocations aimed at minimizing mortality, we observe that at values of *R*_0_ slightly higher than the herd immunity threshold, a discontinuous shift occurs in the optimal allocation for minimizing mortality, towards the higher age groups – see bottom right panel of Figure 4. This shift is associated with a large jump in the number of infections - see top left panel. After this shift occurs, the allocation varies only slightly with *R*_0_, except among age groups 10-19 and 30-39. Intuitively, the above cases are characterized by a significant epidemic outbreak which leads to very high mortality unless vaccines are allocated mostly to those at risk.

The above results show that designing vaccine allocation with the aim of achieving herd immunity is not a robust strategy as it leaves the older age groups exposed, if, due to mis-estimation or to changed circumstances, herd-immunity is not achieved. The results in Figure 4 show that one way to avoid this risk is to opt for vaccine allocations minimizing mortality. This approach, however, leads to a large number of infections, which in turn are associated with an economic price, possible long term public health consequences, and other costs which may offset the benefits in the reduction of mortality. The results presented in Figure 4 show that the trade-offs between infections and the mortality are considerable. For example, when *R*_0_ = 3 and vaccines can be allocated to all ages, mortality ranges from 1.55M to 0.2M, while overall infections range from 75M to 161M, for vaccine allocations aimed at minimizing number of infections, or mortality, respectively. In what follows, we utilize a Pareto optimization approach, which to our knowledge is novel in this context, and which allows one to systematically evaluate the trade-offs involved among the two measures.

### 3.3 Pareto-optimal allocation of vaccines

Section 3.2 considers vaccine allocations that minimize one of two basic measures: attack rate (overall number of infections), and mortality. Due to the age-dependent infection fatality ratio there is a trade-off between infections and mortality, e.g., minimizing mortality by vaccination of the elderly results in more infections of the younger age groups. It is therefore useful to study the problem as one of multi-objective optimization, and therefore apply the notion of Pareto optimality.

It is instructive to first consider a wide set of randomly chosen vaccine allocations, not necessarily designed to be optimal in any sense. For each allocation, we plot the possible outcomes in a plane so that the coordinates of a point correspond to the outcome of a given vaccine allocation in terms of infections and mortality. The yellow points presented in Figure 5A present the outcomes corresponding to allocations with a coverage of 55% of the US population, but restricted to adults of age 20 and over (providing coverage of 73.2% of this group), assuming *R*_0_ = 3 in the post-vaccination period. The point highlighted with a diamond marker, corresponds to a *pro rata* allocation with no prioritization among those aged 20 and over, giving rise to mortality of roughly 750,000 individuals and an overall number of ∼150 million infected individuals. Inspection of the random allocations shows that many alternative allocations achieve better outcomes in both senses, namely reduce both infections and mortality compared to the *pro rata* allocation. Therefore, we consider the curve in the plane of possible outcomes (infections, mortality) which represents the *Pareto front*, the set of outcomes that cannot be improved upon in both senses by changing the allocation, see, e.g., the black solid curve in Figure 5. The choice among outcomes on the Pareto front (and the corresponding vaccine allocations) depends on one’s weighing of the importance of the two measures.

**Figure 5:**
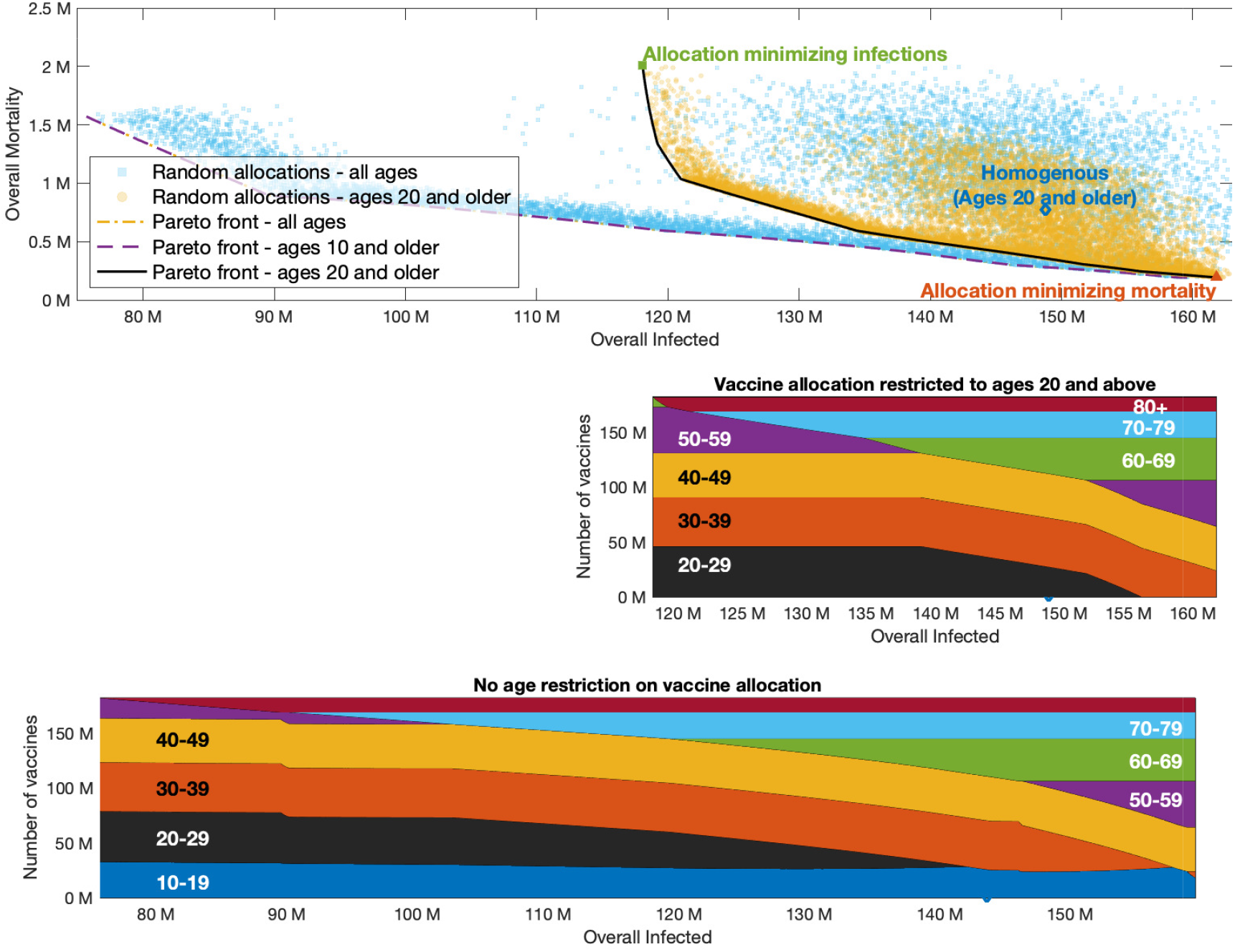
Pareto front. Top graph presented outcomes of random allocations when all ages are eligible for vaccination (square blue markers) and only ages 20 and older are eligible (round yellow markers). Super-imposed are the Pareto fronts in the case ages 20 and older are eligible for vaccination (black solid), ages 10 and older are eligible (yellow dash-dotted) and when all ages are eligible (purple dashed). The latter two curves are indistinguishable. Bottom graphs present vaccine allocation along the Pareto fronts in the case of ages 20 and older are eligible for vaccination, and when all ages or ages 10 and older are eligible for vaccination.

The vaccine allocations corresponding to outcomes along the Pareto front are presented in the bottom panel of Figure 5. The right endpoint of the Pareto front represents the outcome corresponding to an allocation chosen so that mortality is minimized, while the left endpoint represents the outcome of the allocation minimizing infections. As expected, the allocation minimizing infections can be seen to prioritize younger age groups, while the allocation minimizing mortality includes older age groups. Moving along the Pareto front we observe a rather complicated structure of variation in the allocations. For example, we see that the 50-59 age group is included in both the allocation minimizing infections and the allocation minimizing mortality, but it is not included in a wide intermediate range along the Pareto front. We observe that age group 0-9 is not included in the allocations along the Pareto front corresponding to the case in which the entire population is eligible for vaccination. This means that, under the present assumptions, *e*.*g*., that children are 50% less susceptible than adults, it is sub-optimal to vaccinate children under 10, no matter how the two goals (minimizing infection and mortality) are weighed. On the other hand, age group 10-19 is included in the optimal allocation along the entire Pareto front. This implies that restricting vaccination to adults over 20 will worsen outcomes. The extent to which outcomes are degraded by restricting eligibility to those over 20 can be gauged by comparing the two Pareto fronts in the top part of figure 4. Thus, for example, if vaccines are restricted to the 20+ age group, then (for *R*_0_ = 3 and when an overall of 55% of the population is vaccinated) the minimal number of infections that can be achieved is 118 million, and the allocation achieving this outcome would given rise to a mortality of 2 million. If those of age 10-19 become eligible, and an allocation generating the same number of infections is chosen, mortality is reduced to 0.6 million.

### 3.4 Effect of assumptions on the relative susceptibility of children

Our baseline examples, presented in Figures 2, 3A, 4 and 5, all adopt the age dependent susceptibility profile estimated in [9], in which the relative susceptibility of age group 0-19 is roughly half that of older age groups. We now consider the impact of changes in this assumption on the optimal allocation. We note that we have also considered the impact of changes in other assumptions concerning, e.g,. vaccine efficacy or vaccine hesitancy, and have found the optimal allocations presented in Sections 3.1-3.3 are quite robust to such changes. Namely, while epidemic outcomes may vary due to changes in assumptions, we found that the optimal allocations do not change as much. See appendix B for additional details.

Let us consider a modified age-dependent susceptibility profile in which adolescents (age group 10-19) are equally susceptible as adults, and an additional modified profile in which all children (age group 0-19) are equally susceptible as adults.

Figure 6 presents the vaccination coverage required for herd immunity and the vaccine allocations that lead to herd immunity at minimal vaccination coverage under the two alternative assumptions on children’s susceptibility to infection. As expected, when the susceptibility of adolescents is higher than the susceptibility of younger children, the allocations designed to achieve herd immunity at minimal coverage dedicate larger portions of vaccines to age group 10-19 at the expense of vaccine allocation in age group 0-9. Herd immunity can be achieved without vaccinating children of ages 0-9 for *R*_0_ up to 4.2.

**Figure 6:**
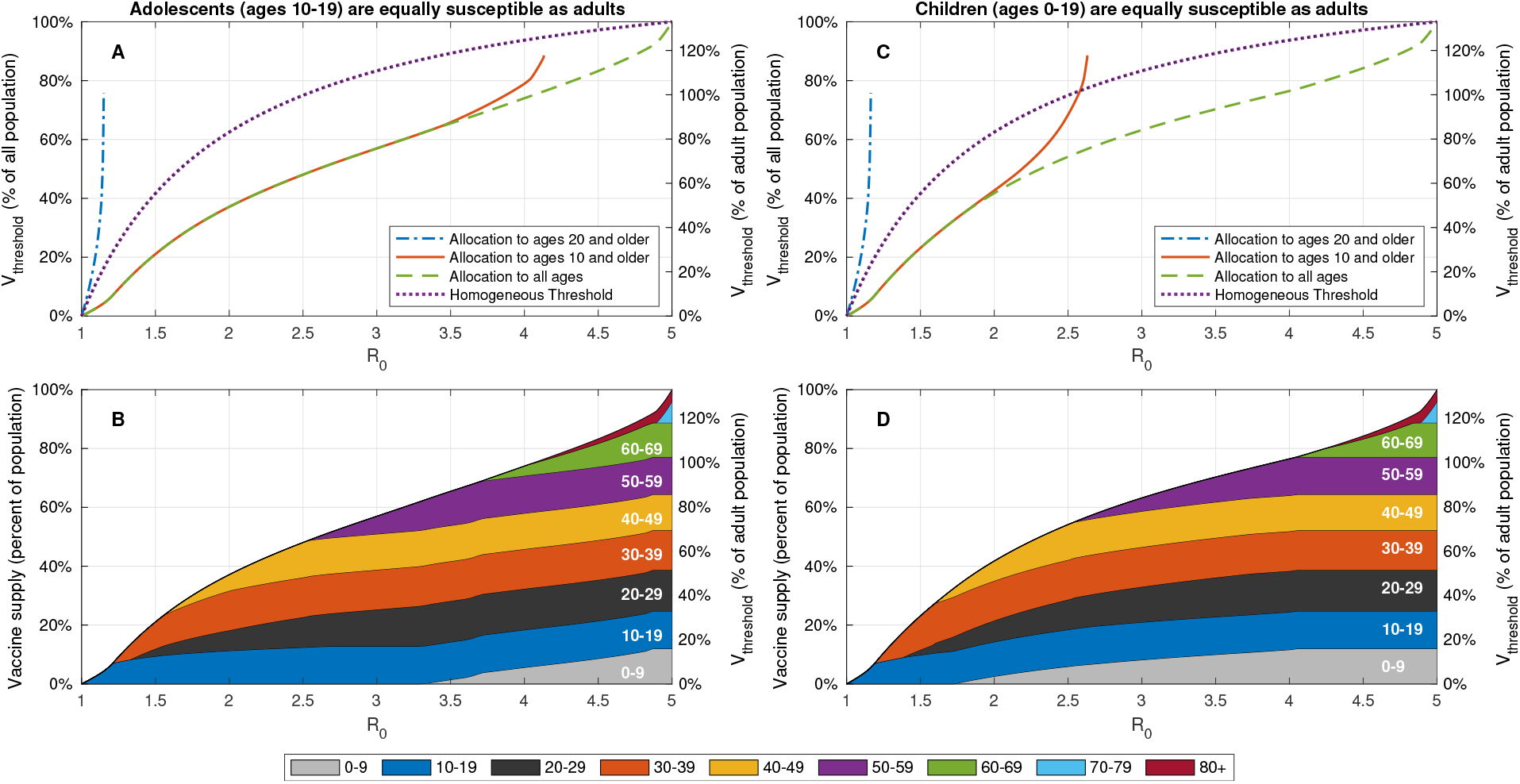
Effect of change in childrens’ susceptibility on vaccination coverage required for herd immunity. A,C: Vaccine coverage *V*_threshold_ required to achieve herd immunity threshold as a function of the reproduction number *R*_0_ for the USA demography and contact structure. The gray curves correspond to the case of 80% vaccine efficacy. B,D: Vaccine allocations at which herd immunity is achieved at minimal vaccine coverage and when there is no age restriction on vaccine allocation.

If we assume that all children are equally susceptible as adults, vaccination of age group 10-19, and to a lesser extent, the vaccination of age group 0-9 becomes of higher priority - indeed in this case herd immunity cannot be reached without vaccinating children in age group 0-9 for *R*_0_ *>* 2.6.

Figures 3B and 3C show similar behavior for other countries.

Examining the outcomes associated with optimal vaccination when herd immunity cannot be achieved, we find that, as expected, when the susceptibility of children is higher, the allocations minimizing infections dedicate larger portions of vaccines to age group 0-19, see Figures 7 and 8. Nevertheless, the allocations minimizing mortality do not change.

**Figure 7:**
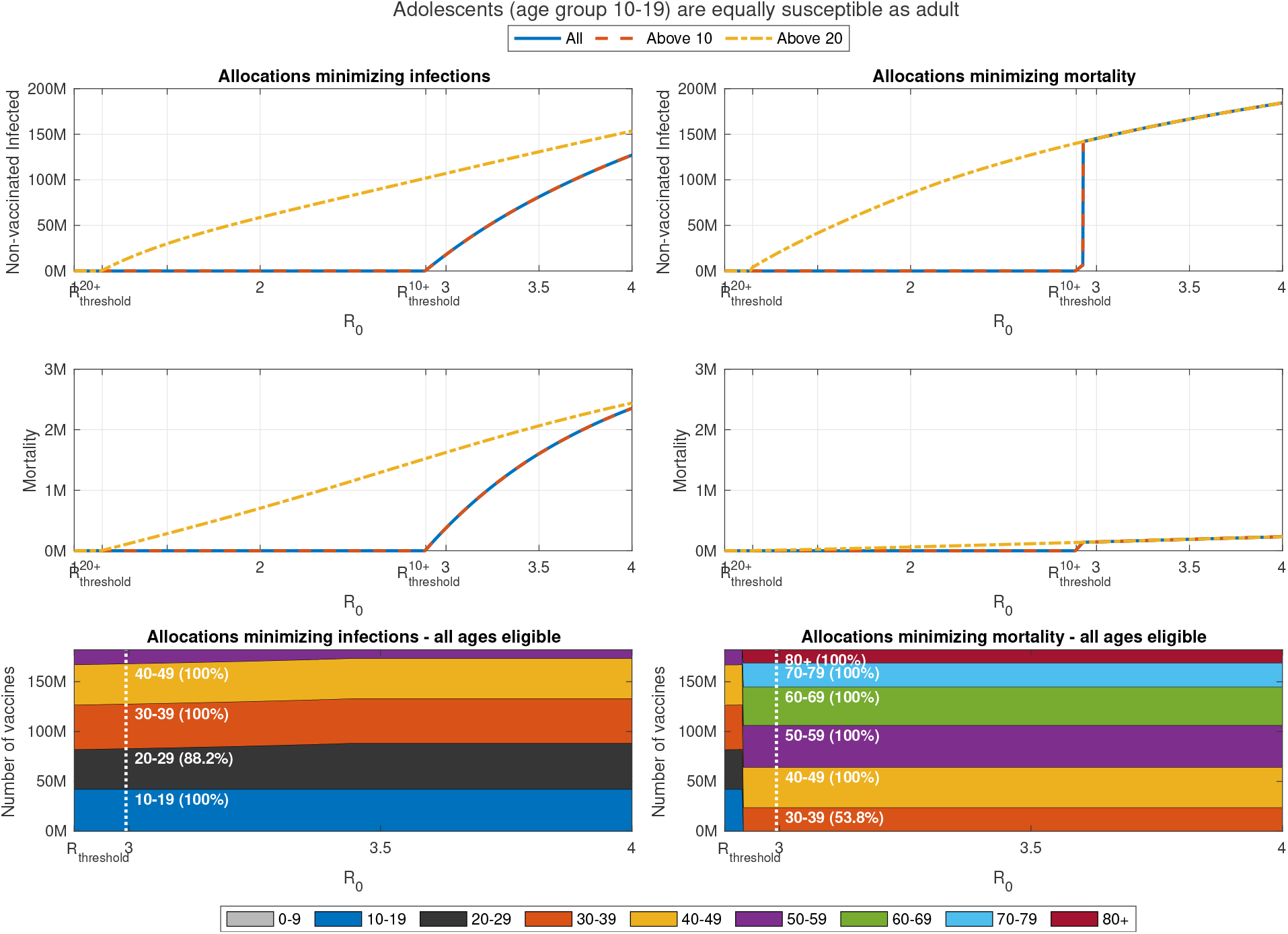
Impact of change in reproduction number when adolescents (age group 10-19) are equally susceptible as adults. Same as Figure 5 except the susceptibility of age group 10-19 is increased by factor of 2 to that of older age groups. Susceptibility of age group 0-9 is not modified.

**Figure 8:**
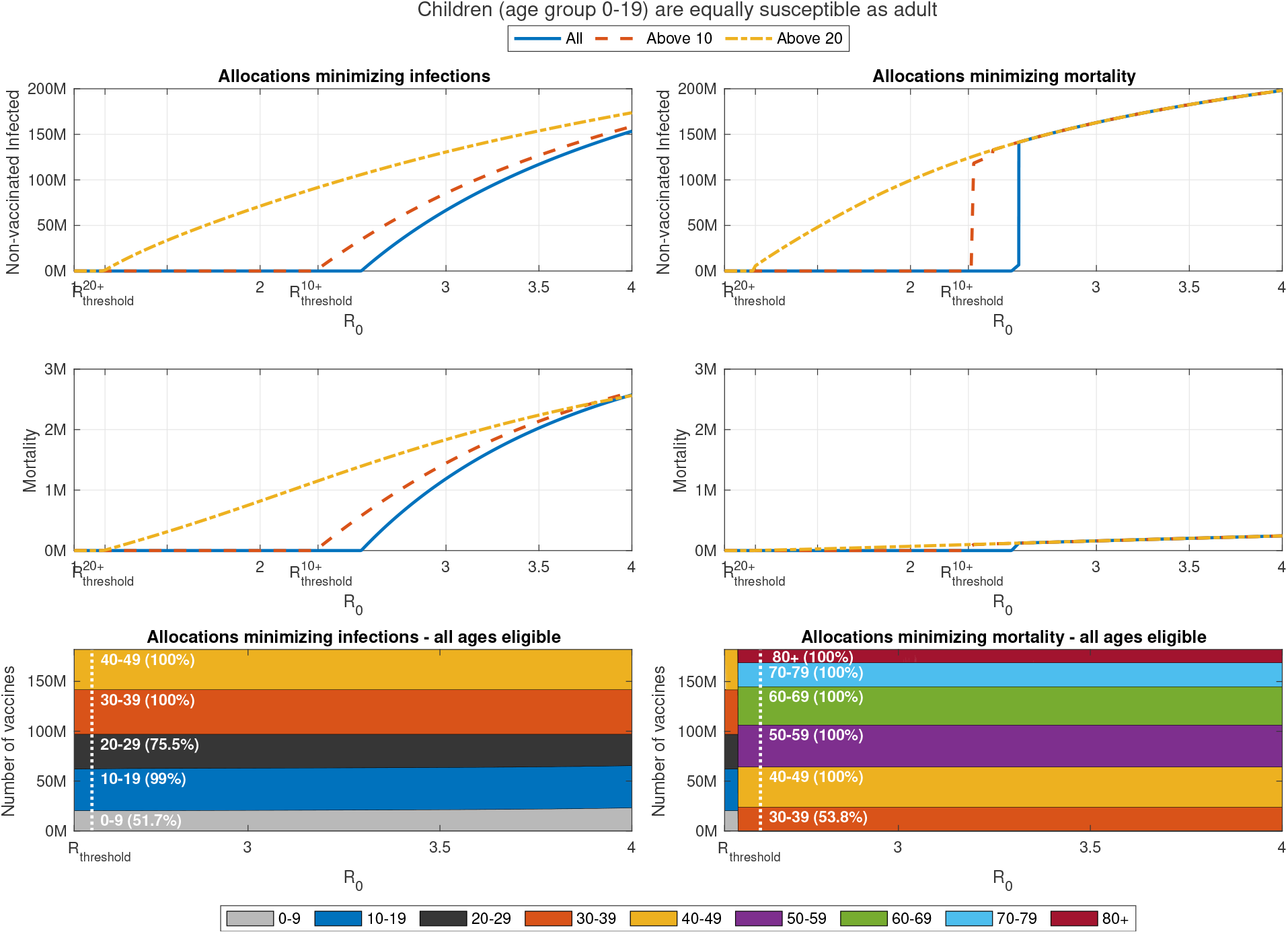
Impact of change in reproduction number when when children (age group 0-19) are equally susceptible as adults. Same as Figure 5 except the susceptibility of age group 0-19 is increases by factor of 2 to that of older age groups.

## 4 Discussion

This study uses modelling and optimization to explore the outcomes of vaccination campaigns for SARS-CoV-19, with emphasis on the effects of vaccinating children. We demonstrate the use of Pareto front computations to systematically evaluate the trade-offs involved among conflicting measures for optimizing vaccine allocations such as mortality and attack rate. In particular, we utilize this approach to compare optimal achievable outcomes when all age-groups can be vaccinated to those that can be attained when younger age groups are not eligible for vaccination.

Our study focused on two questions:

- How essential is the vaccination of children and youths to achieving herd immunity?
- What is the population level impact of vaccination of children in case herd immunity by vaccination cannot be attained?

Regarding the first question, the results presented in section 3.1 show that, for a wide range of values of the reproductive number *R*_0_, a ‘herd immunity’ strategy is not feasible if vaccination is restricted to ages 20 and older. Indeed, in this case, even if the entire eligible population is vaccinated, the maximal basic reproduction number at which herd immunity can be achieved is around 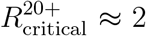 and increases only slightly when taking into account existing immunity due to prior infections. When ages 10 and older are eligible for vaccination, optimal allocations can lead to herd immunity for basic reproduction numbers up to 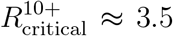, though this threshold varies significantly (±20%), e.g., across countries and in dependence on vaccine efficacy and prior immunity. Interestingly, the above thresholds are only weakly correlated with the percentage of children in the population and rather depend on the level of assortativity of mixing within the children sub-population, as reflected in the contact matrix.

Our key results in section 3.2 indicate that

- Designing vaccine allocation with the aim of achieving herd immunity is not a robust strategy as it leaves the older age groups exposed, if, due to a mis-estimation or to changed circumstances, herd-immunity is not achieved.

For example, though herd immunity can be achieved at the range 2.5 − 3.5 of basic reproduction number estimated for wild-type of SARS-CoV-19 [29] and perhaps for the approximately 50% higher range of values estimated for the Alpha variant [30], it cannot be achieved solely by vaccination at the estimated basic reproduction number of the Delta variant [31]. In case allocations are optimized to achieve herd immunity and *R*_0_ increases to a sufficiently high level so that herd immunity is not achieved, mortality would rise steeply since these allocations give preference to the young and leave the older age groups exposed. Thus, even if according to current estimates, the ‘herd immunity’ (through vaccination) strategy is feasible and is therefore theoretically the optimal strategy, it may not be the strategy of choice given the risks involved in mis-estimation of parameters or due to a genuine increase in *R*_0_.

The second question we posed concerns the cases in which the ‘herd immunity’ strategy is not feasible. Our key findings in this case are

- Vaccine allocations aimed solely at minimizing mortality are only marginally affected by the ineligibility of age groups 0-9 or 0-19. These allocations, however, give rise to a large number of infections with a high attack rate in children.
- Optimal vaccine allocations that also give weight to the number of infections include age group 10-19. Consequently, ineligibility of age group 10-19 results in worse outcomes. On the other hand, age group 0-9 is not included in these optimal vaccine allocations. These results suggest that, under the criteria studied, vaccination of age group 0-9 should not be of priority.

Indeed, when emphasis is given to minimizing mortality, in all scenarios examined, we observed that vaccines are allocated to the entire population (up to constraints related to hesitancy) in age groups over 60, and no vaccines are allocated to the age group under 10. In some cases, vaccines were allocated to age group 10-19 but in these cases we observed that the mortality and infections are only marginally reduced relative to allocations which do not include age group 10-19. The ‘protective’ allocations, however, are associated with a large number of infections. In the example of the USA with 55% vaccination coverage and *R*_0_ = 3, these allocations result in the infection of roughly 161 million people (47% of the population), mostly from age group 0-19. While the severity of COVID-19 in children is typically very low, such large numbers may carry short or longer-term risks [4].

When considering allocations of vaccines to minimize the number of infections, one might expect that vaccination of children and adolescents would play a greater role than in the case of minimizing mortality, in view of the fact that mortality minimizing allocations seek to protect the older, more vulnerable age groups. This expectation is borne out in the case of age group 10-19, which in the optimal allocation for minimizing the number of infections receives a high coverage. However, we also find that the age group 0-9 is either not included at all in this allocation, or, for higher values of *R*_0_, included with a very low coverage. It should be noted that when emphasis is given to minimizing infections, we observe that the infections-minimizing allocation give rise to high mortality.

The two strategies discussed above reflect two extremes - prioritization of younger age groups to maximize indirect protection (minimize infections) vs prioritization of older age groups to maximize direct protection (minimize mortality). In this work, we go beyond these two extremes and present a spectrum of Pareto-optimal strategies that give varying weights to both indirect and direct protection. We show, in the USA example, that, along the spectrum of strategies, the number of infections varies in the range of roughly 75-161 million people (22-47% of the population). Thus, the trade-off between infections and mortality is substantial. Application of the Pareto front allows one to systematically evaluate these trade-offs. The Pareto front allows one to make an informed choice of the allocation policy. For example, one might choose an allocation whose estimated cost with respect to mortality is 10% higher than the minimal mortality possible in order to achieve a a 25% reduction in the number of infections.

In our basic scenario we assumed that the relative susceptibility of age group 0-19 is half that of adults [9]. As we have shown, if only age group 0-9 has lower susceptibility while age group age group 10-19 is as susceptible as adults, the previous conclusions about the importance of vaccination of the 10-19 age group and the fact that vaccination of the 0-9 group is non-optimal become even stronger.

We should stress here that the above results should not be taken to imply that vaccination of children is not in itself beneficial. The comparisons here are performed under the assumption of a fixed supply of vaccines, in which case vaccination of one age group involves an opportunity cost in not vaccinating another. Obviously if one can extend the coverage so as to include children, without reducing vaccination levels in other age groups, then doing so will only improve outcomes.

This study is subject to several limitations. We focus on the outcomes in the medium-term range after the vaccination campaign has ended. Over longer timescales, the possibilities of waning immunity and virus mutation might influence these predictions. Our study optimizes outcomes for the post-vaccination phase, and is, therefore, most relevant when disease spread is contained during the vaccination campaign, e.g., by non-pharmaceutical interventions. In this case, once a vaccine allocation that is optimized for post-vaccination outcomes is determined, transient features of a vaccination campaign that results in the desired allocation can be designed, for example, to allow gradually relaxing non-pharmaceutical interventions during the campaign [15]. Accordingly, we do not account for increased mortality rates during periods of excessive hospital load [32]. In case the vaccination campaign occurs in parallel to an ongoing outbreak, short-term goals are likely to dominate the design of the vaccination campaign [16]. We have also used pre-pandemic contact matrices in accordance with the aim of returning to pre-pandemic routine after the vaccination campaign. Nevertheless, we present results for a range of basic reproduction numbers *R*_0_, and therefore implicitly account for a new routine which includes a degree of non-pharmaceutical interventions. Long-term changes in school operation, however, are not well captured by this approach. Accounting for such age-dependent non-pharmaceutical interventions will require the estimation and application of post-pandemic contact matrices.

The results presented in this study, as well as results in other settings, may be further explored using open-source tools that accompany this study. These tools can also be applied at various stages of an vaccination campaign to optimize the allocation of the remaining available vaccines according to current estimates, taking into account those already vaccinated. An application of these tools to real world data will be presented elsewhere.

The debate over childrens’ vaccination is on-going in many countries, and will likely reemerge in others as children younger than 12 also become eligible for vaccination. We believe that the approach presented in this work can provide valuable model-informed tools to assist decision making in these matters.

## Data Availability

This work is based on the use of public aggregate data, which is fully available in the codes provided.

https://github.com/NGavish/OptimalVaccinationRoleOfChildren

## Acknowledgments

This research was supported by the ISRAEL SCIENCE FOUNDATION (grant No. 3730/20) within the KillCorona – Curbing Coronavirus Research Program. We are grateful to Dr. Amit Huppert for discussions and valuable comments.

## Supplementary material

### A Codes

The codes used to produce the graphs in this work are available in https://github.com/NGavish/OptimalVaccinationRoleOfChildren

### B Impact of changes in parameters

We consider the impact of changes in assumptions concerning, e.g,. vaccine efficacy or vaccine hesitancy. The effects of assumptions on the relative susceptibility of children were discussed in Section 3.4.

In what follows, we consider the impact of such changes either at a given *R*_0_ or for a range of basic reproduction number. We note that in Section 3.4 we avoided studying the effect of assumptions on the relative susceptibility of children by considering epidemic outcomes at the same basic reproduction number. The reason is that the formula defining the basic reproduction number depends on the susceptibility profile, see Eq. 7. Namely, the value of the basic reproduction number changes as a result of changes in assumptions on relative susceptibility.

### B.1 Results for all-or-none vaccines

We allow for the possibility that a fraction 1 − *ν* of the population vaccinated does not generate immunity, while the rest of the vaccinated population is fully immune (*ε* = 0). This case is known as ‘all-or-none’ vaccine, whereas the case in which all the vaccinated population is partial immune (0 *< ε <* 1) corresponds to the case of ‘leaky’ vaccines. The computation of vaccine supply thresholds and the critical reproduction numbers are indifferent to the ‘leaky’ or ‘all-or-none’ nature of vaccine protection. Indeed, these computations rely on the absolute value of the dominant eigenvalue of (7), and thus their dependence on *ν* and *ε* is only through the expression *ϵν*.

Leaky vaccines are known to result in a higher prevalence of infection than ‘all-or-none’ vaccines [33]. Indeed, for the USA example presented in Figure 5, we observe that allocations along the Pareto front corresponding to ‘all-or-none’ vaccines give rise to 20% less infections than allocations along the Pareto front corresponding to ‘leaky’ vaccines, see Figure 9. Details of the outcomes of a vaccination campaign with ‘all-or-none’ vaccines are presented in Figure 10. In cases A and B of Figure 10, the reductions in overall infections are 28% and 25%, respectively. Finally, we study the impact of vaccine nature on optimal allocation as the reproduction number changes. We observe only a change in the allocation minimizing mortality concerning the allocation of vaccine to the 10-19 or 30-39 age groups.

**Figure 9:**
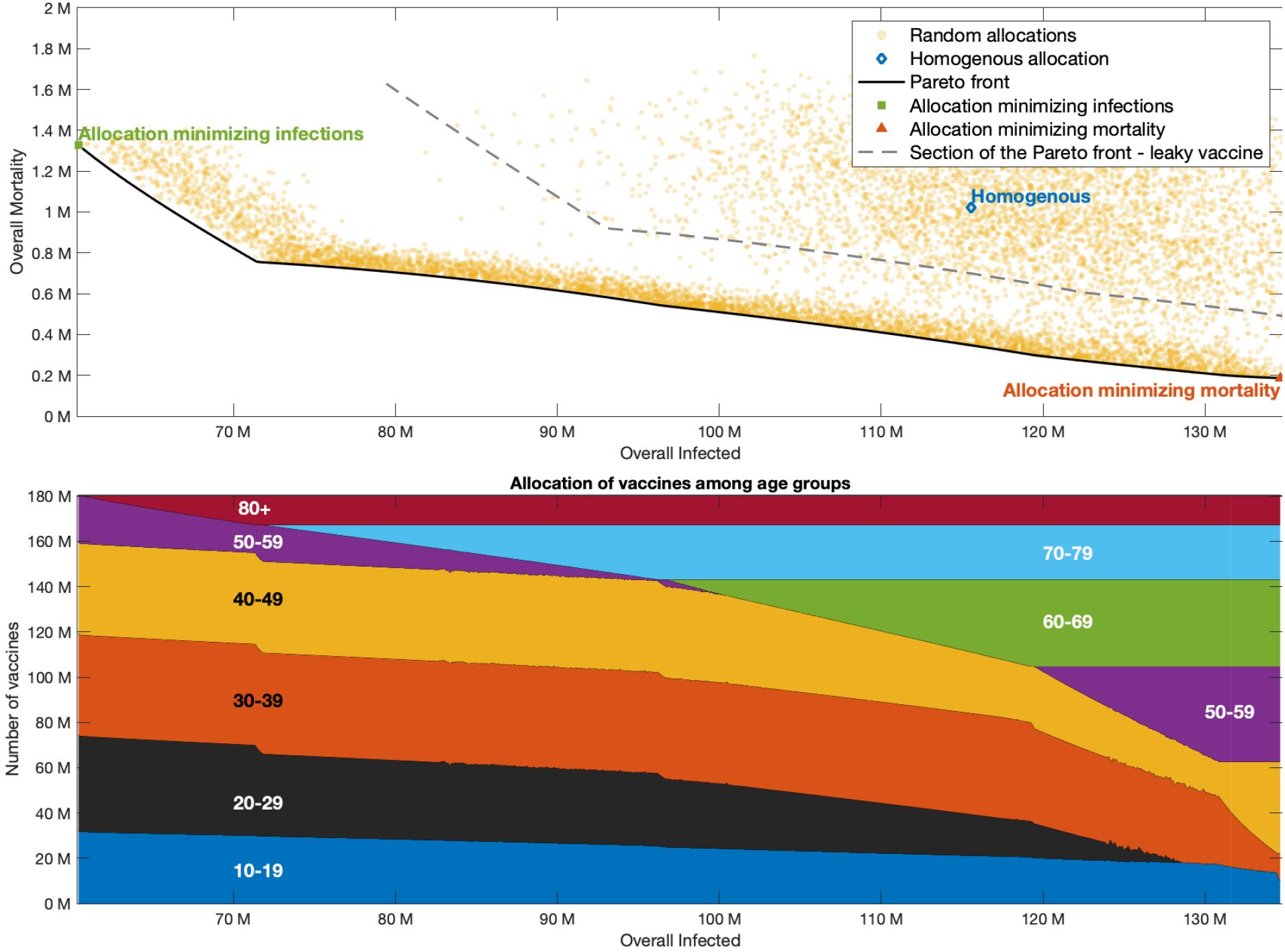
Pareto front with all-or-none vaccine. Same as Figure 5, but with 80% all-or-none vaccines. The gray curves in the upper plot correspond to the case of leaky vaccines.

**Figure 10:**
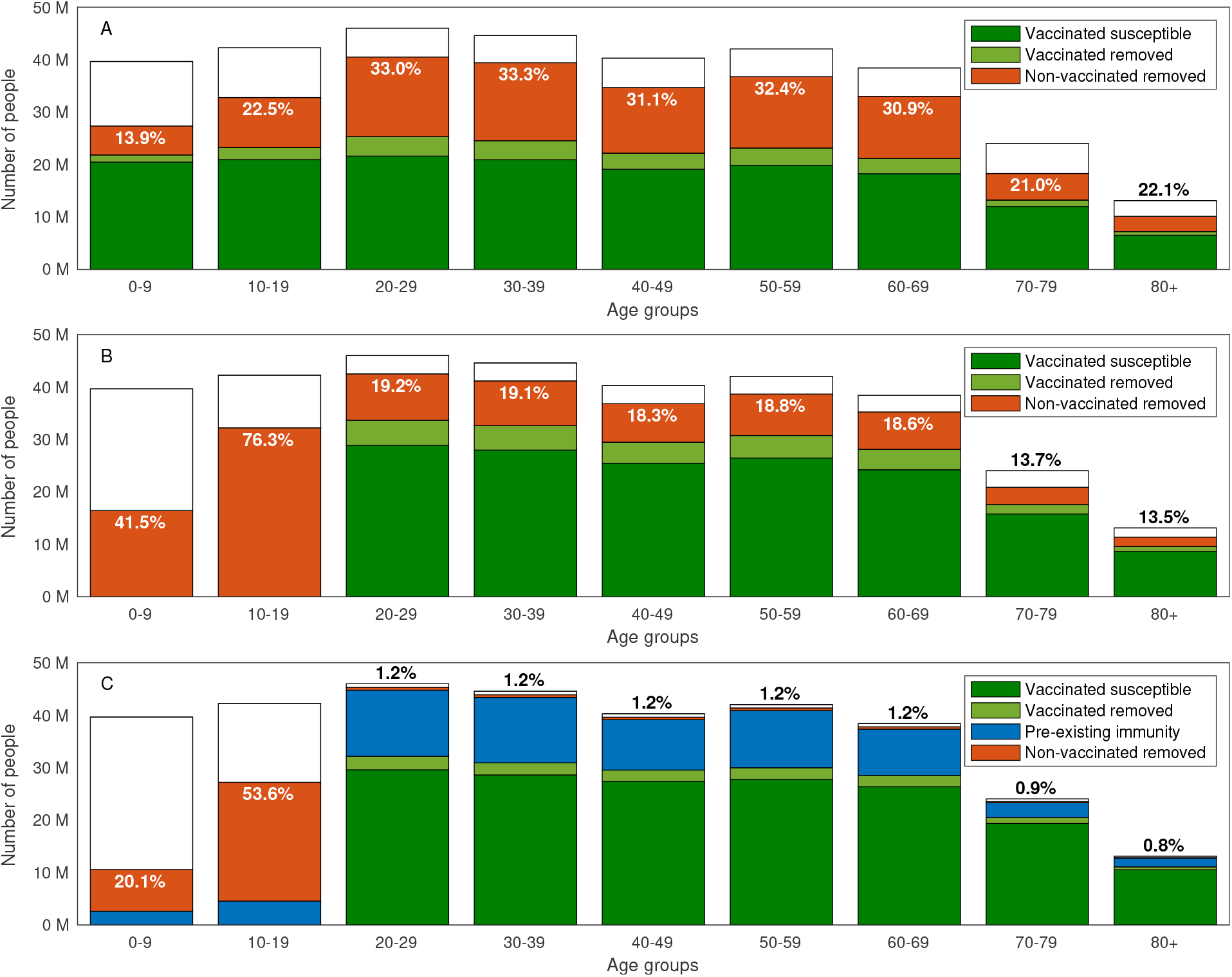
Final size of epidemic with all-or-none vaccine. Same as Figure 1 but with all-or-none vaccines: The expected outcome of a partial return to normality in the USA to a basic reproduction number of *R*_0_ = 3 after completion of a vaccination campaign covering 55% of the population. Removed population refers to those recovered or dead. The computation considers a vaccination campaign in which A: All the population is eligible for vaccinations. B: Vaccine eligibility is limited to ages 20 and above. C: Vaccine eligibility is limited to ages 20 and above, and at the time normality is restored 20% of the population is recovered from COVID-19, and the prevalence of active cases is 0.5% of the population. The text in all graphs corresponds to the percent of non-vaccinated removed individuals in each age group.

### B.2 Impact of vaccine efficacy

We consider the impact of vaccine efficacy in reducing the susceptibility of those vaccinated on the outcomes of the vaccination campaign.

The relative susceptibility *ε* of vaccinated individuals effects the vaccine supply thresholds and the critical reproduction numbers solely through the expression *εp*_*i*_ in (7), were *p*_*i*_ is the portion of age group *i* that is vaccinated. Therefore, in terms of vaccine supply thresholds and the critical reproduction numbers, changes in vaccine efficacy are equivalent to changes in vaccination coverage. As expected, increase of vaccine efficacy lowers the vaccine supply threshold required for herd immunity and vice-versa, see Figure 12. Particularly, children under the age of 10 appear in the allocation that achieves herd immunity with minimal converge at higher values of *R*_0_. Similar behavior is observed in computations adapted for various countries.

**Figure 11:**
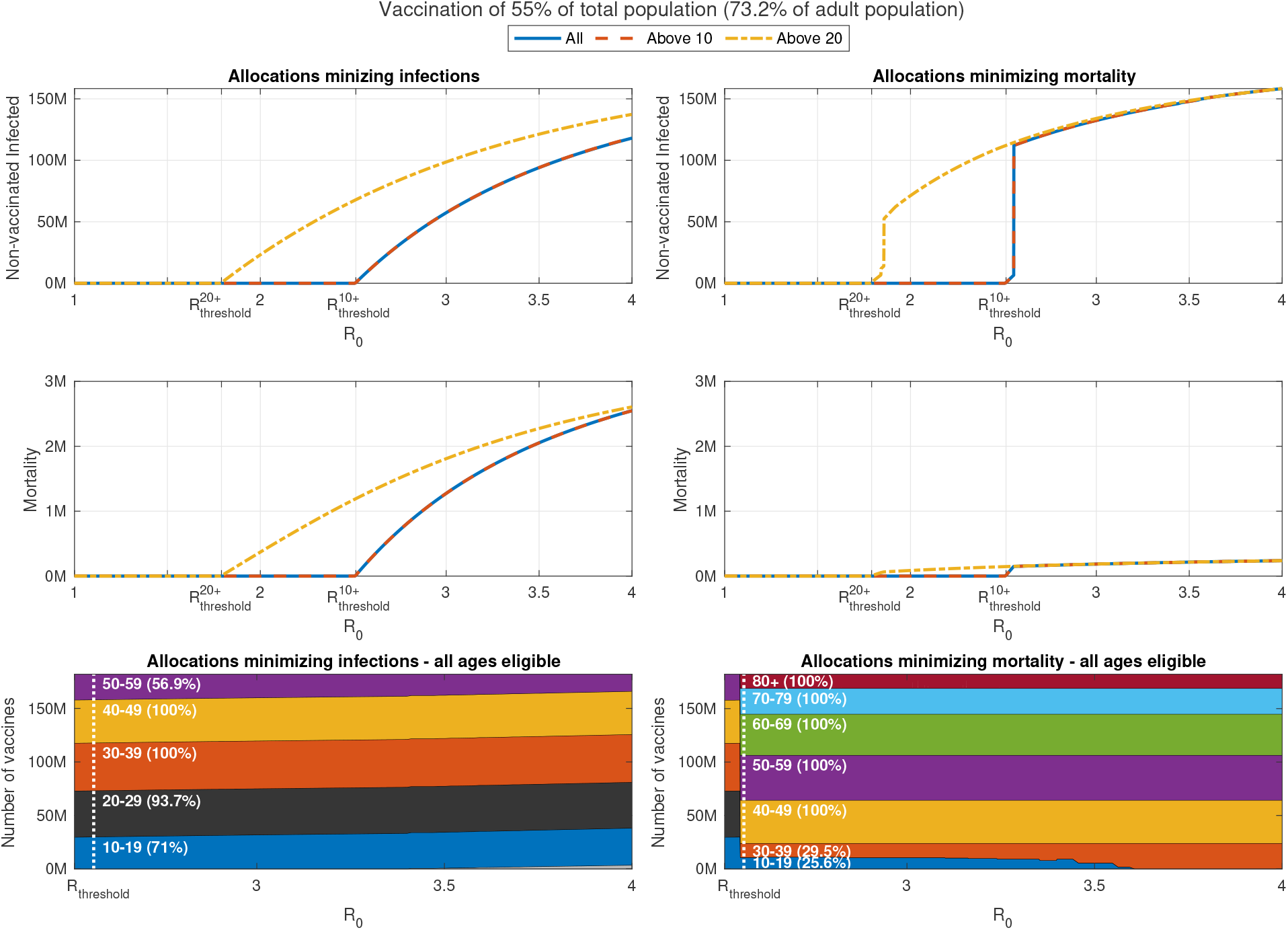
Impact of change in reproduction number with all-or-none vaccines. Same as Figure 4, but with 80% all-or-none vaccines.

**Figure 12:**
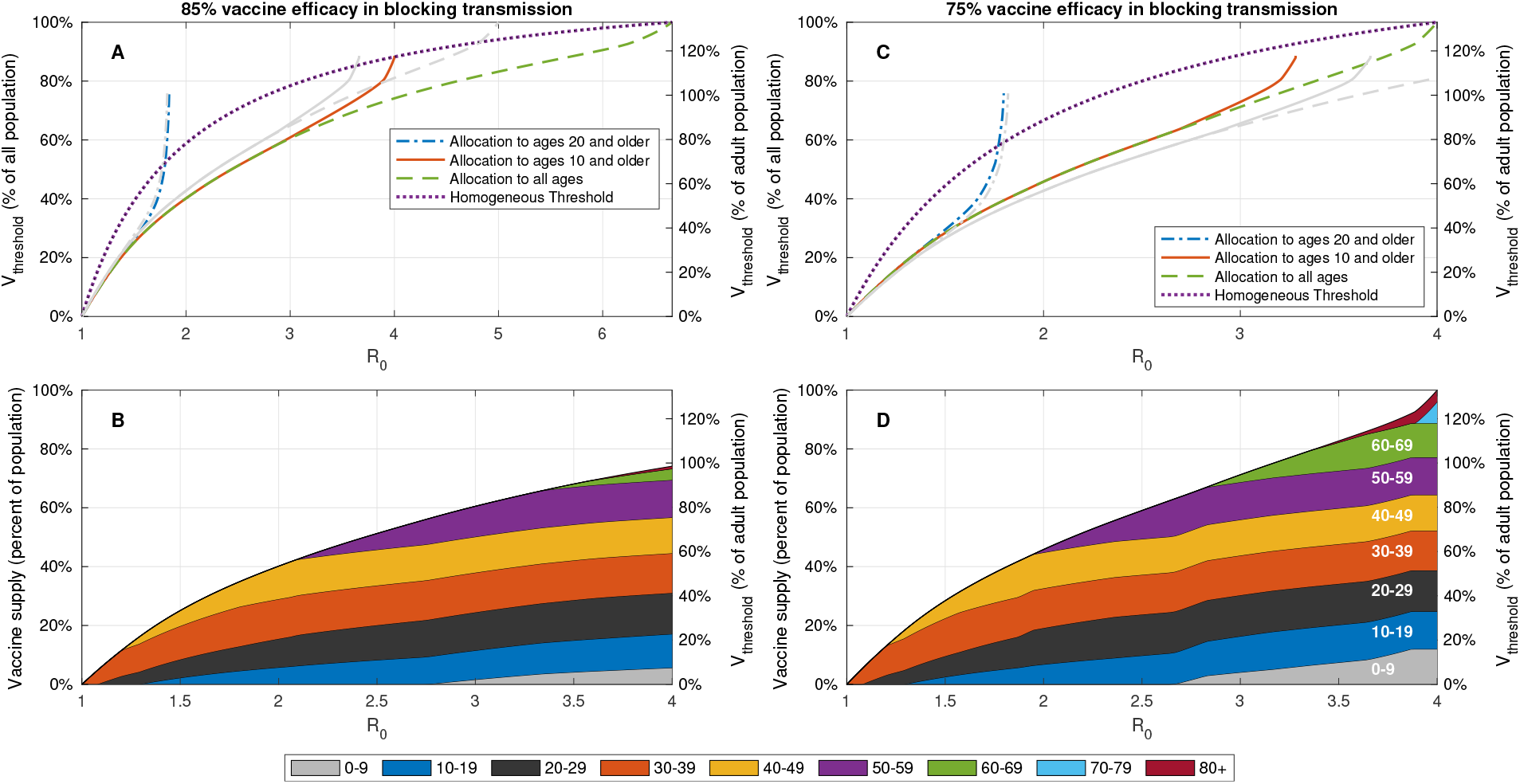
Effect of vaccine efficacy in blocking transmission (reducing susceptibility) on vaccination coverage required for herd immunity. A,C: Vaccine coverage *V*_threshold_ required to achieve herd immunity threshold as a function of the reproduction number *R*_0_ for the USA demography and contact structure. The gray curves correspond to the case of 80% vaccine efficacy. B,D: Vaccine allocations at which herd immunity is achieved at minimal vaccine coverage and when all the population is eligible for vaccination.

Particularly, we find that an change of 5% in vaccine efficacy shifted the critical reproduction number 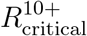 by Δ*R*_0_ = 0.25 on average, see Figure 13. However, when only ages 20 and older are eligible for vaccination, the shift in the critical reproduction number 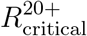 is much smaller.

**Figure 13:**
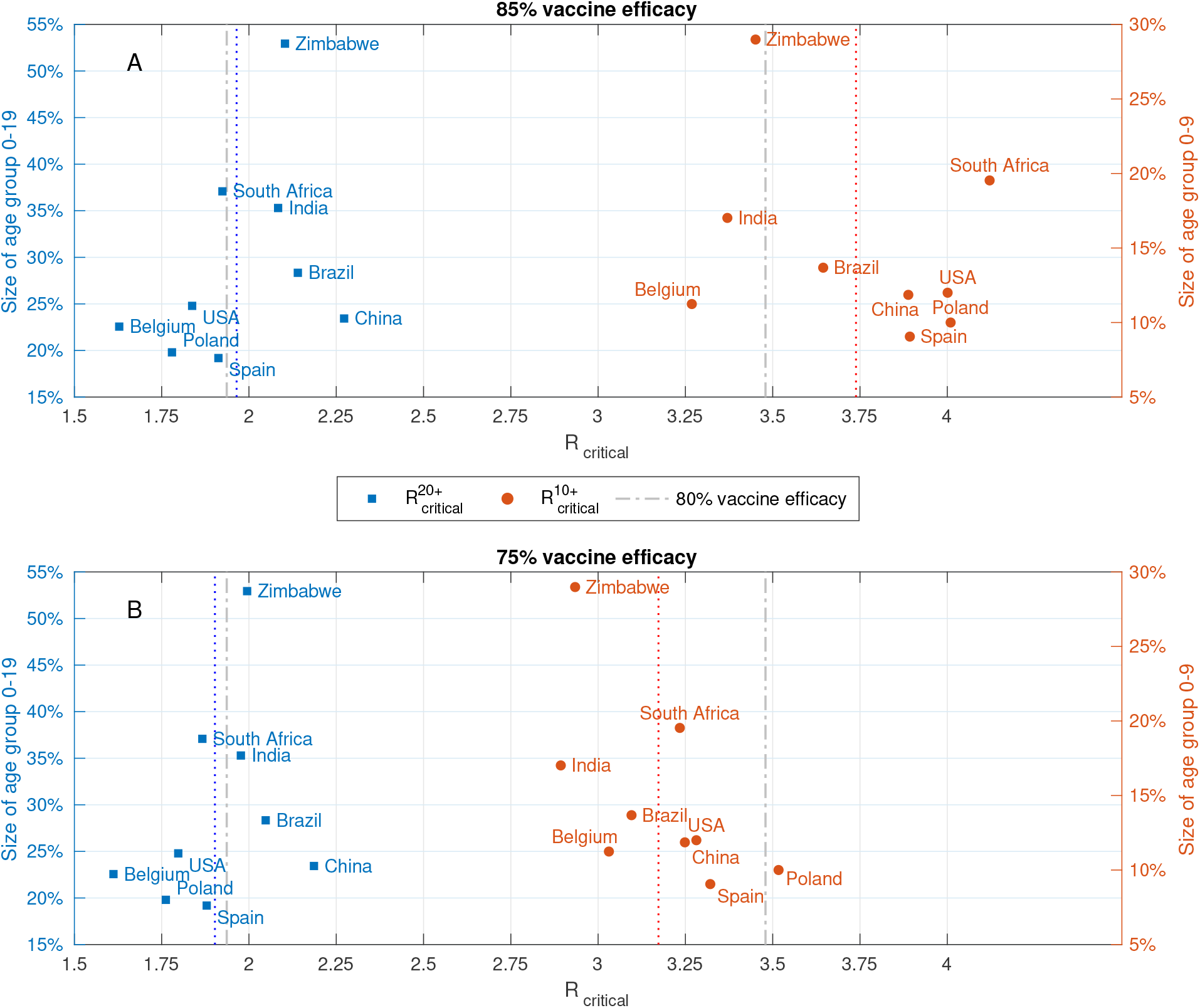
Effect of vaccine efficacy on critical reproduction numbers. Reproduction numbers 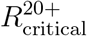 and 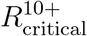 at which herd immunity cannot be achieved without vaccination of age groups 0 − 19 and 0 − 9, respectively. Computed for A: 85% vaccine efficacy in blocking transmission. B: 75% vaccine efficacy in blocking transmission. The gray curves in both plots correspond to the case of 80% vaccine efficacy.

We further consider the impact of vaccine efficacy in cases in which herd immunity is not achieved. As expected, the optimal outcomes that result from optimal vaccine distribution along the Pareto front improve with an increase in vaccine efficacy. We observe that a 5% change in vaccine efficacy leads to roughly 33% change in the minimal overall mortality that can be achieved, and 40%-70% change in the minimal overall infections that can achieved by proper vaccine allocation, see Figure 14. Moreover, we observe that as vaccine efficacy decreases, the optimal vaccine allocation shifts toward the vaccination of younger ages.

**Figure 14:**
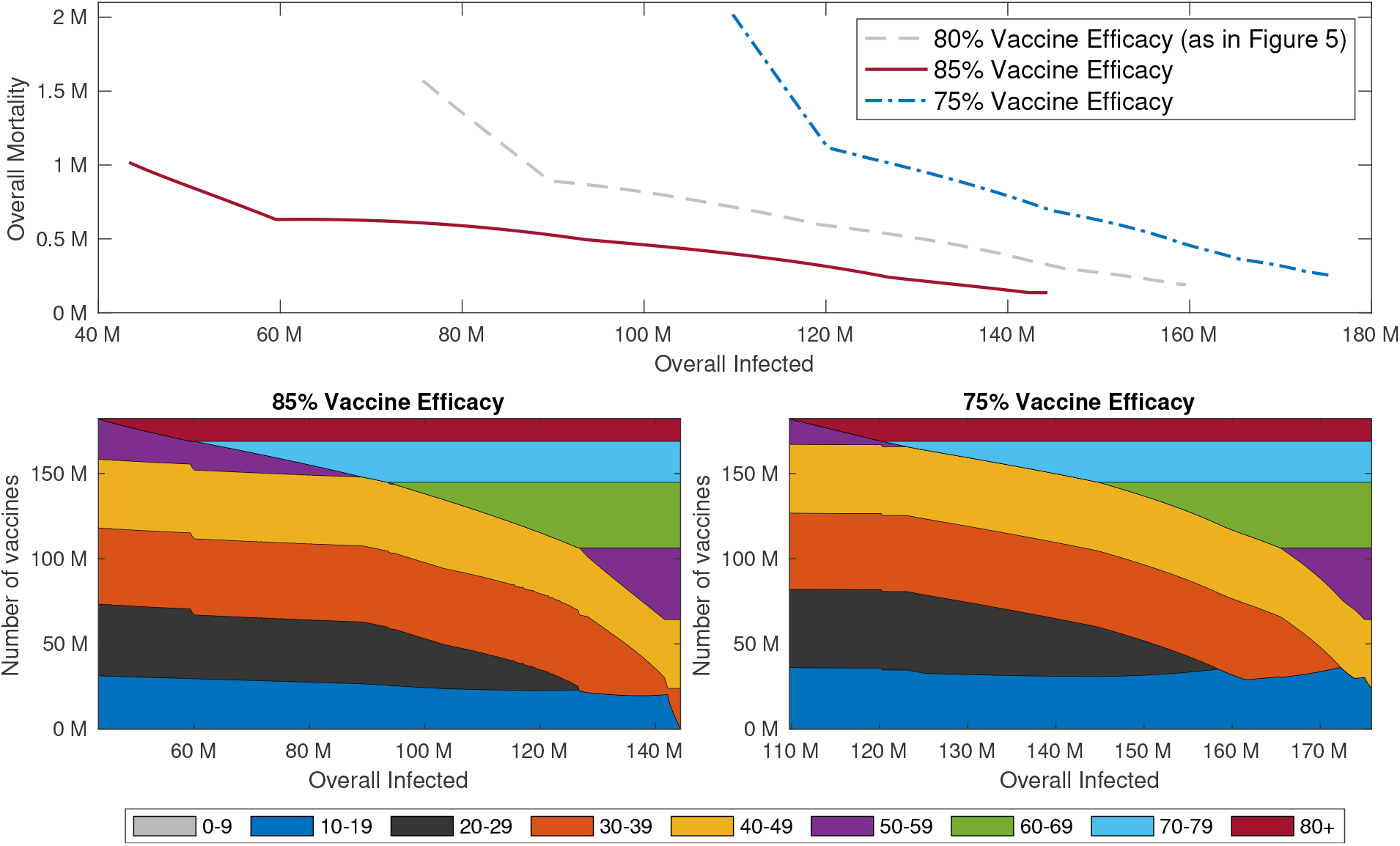
Pareto fronts for different vaccine efficacy. Top graphs: Pareto fronts corresponding to 85% (solid red), 80% (dashed gray) and 75% vaccine efficacy (dash-dotted blue). All other parameters are as in Figure 1. Bottom Graphs: Vaccine allocations along the corresponding Pareto fronts with 85% vaccine efficacy (left) and 75% vaccine efficacy (right).

### B.3 Impact of vaccine coverage

When herd immunity is not achievable, vaccine coverage becomes a key parameter in the design of a vaccination campaign. As expected, the optimal outcomes that result from optimal vaccine distribution along the Pareto front improve with an increase in vaccine efficacy. We observe that a 5% change in vaccine coverage leads to roughly 20% change in the minimal overall mortality that can be achieved, and 50% change in the minimal number of infections that can achieved by proper allocation of vaccines, see Figure 15. The reason for the relatively small change in overall mortality is that already at relatively low vaccine coverage the optimal vaccine allocation for minimizing mortality spans the older age groups, and additional vaccine coverage is mostly utilized to extend allocations to younger age groups which provide indirect protection.

**Figure 15:**
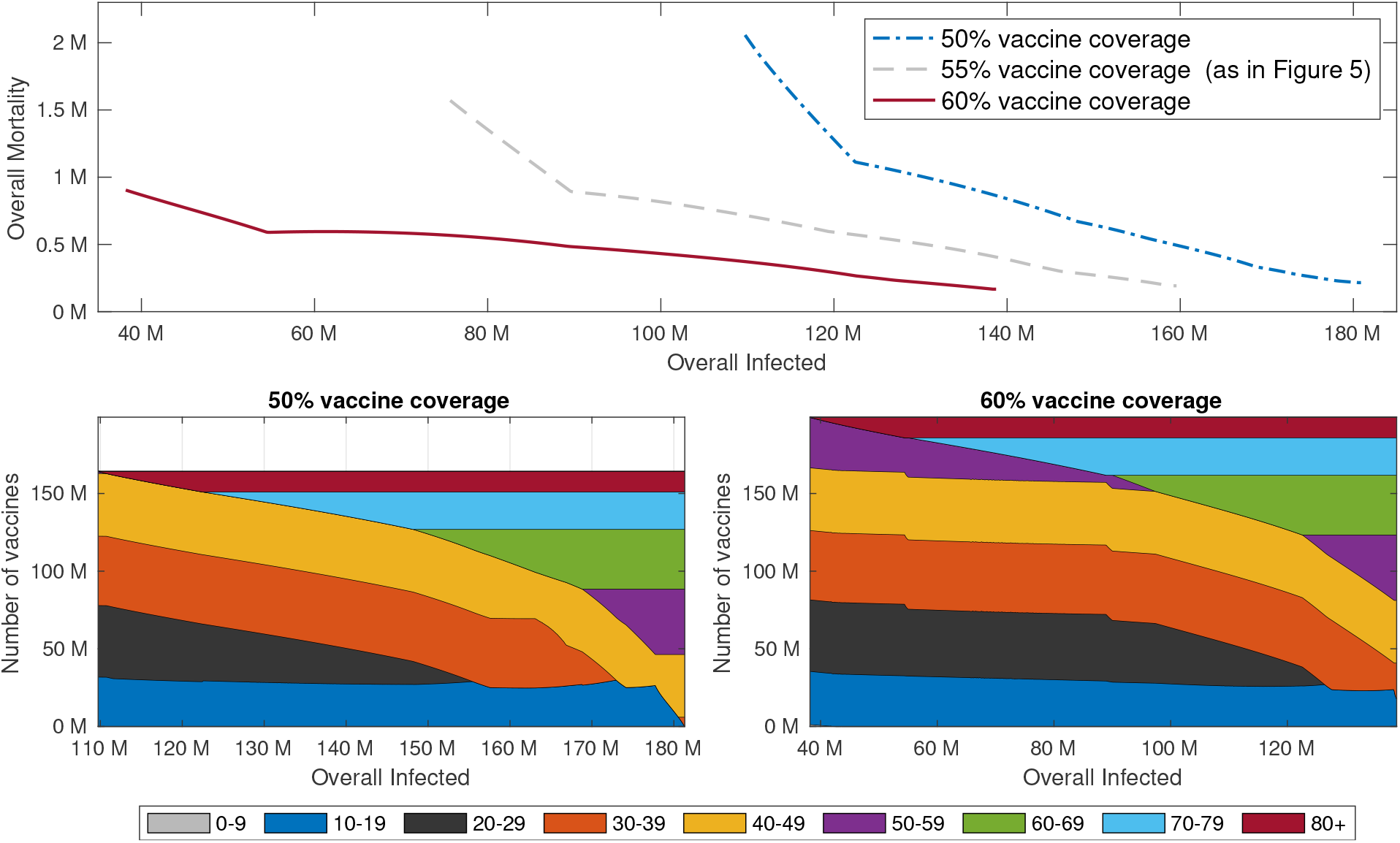
Pareto fronts for different vaccine coverage. Top graphs: Pareto fronts corresponding to 50% (solid red), 55% (dashed gray) and 60% vaccine coverage (dash-dotted blue). All other parameters are as in Figure 5. Bottom Graphs: Vaccine allocations along the corresponding Pareto fronts with 50% vaccine coverage (left) and 60% vaccine coverage (right).

### B.4 Impact of vaccine hesitancy

The examples presented in Figures 1-5 all consider cases where vaccine allocations can include vaccination of 100% of an age group. However, vaccine hesitancy, logistical difficulties and existing medical conditions are likely to limit actual vaccination coverage within an age group. The constraint that vaccine allocation cannot exceed 90% of each age group leads to a significant reduction in the critical reproduction number 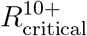 for which herd immunity is achievable by proper allocation of vaccines from an average of 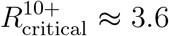 in various countries to 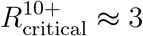, see Figure 16. Imposing a stricter constraint that vaccine allocation cannot exceed 80% of each age group, leads to a further reduction to 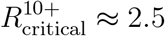.

**Figure 16:**
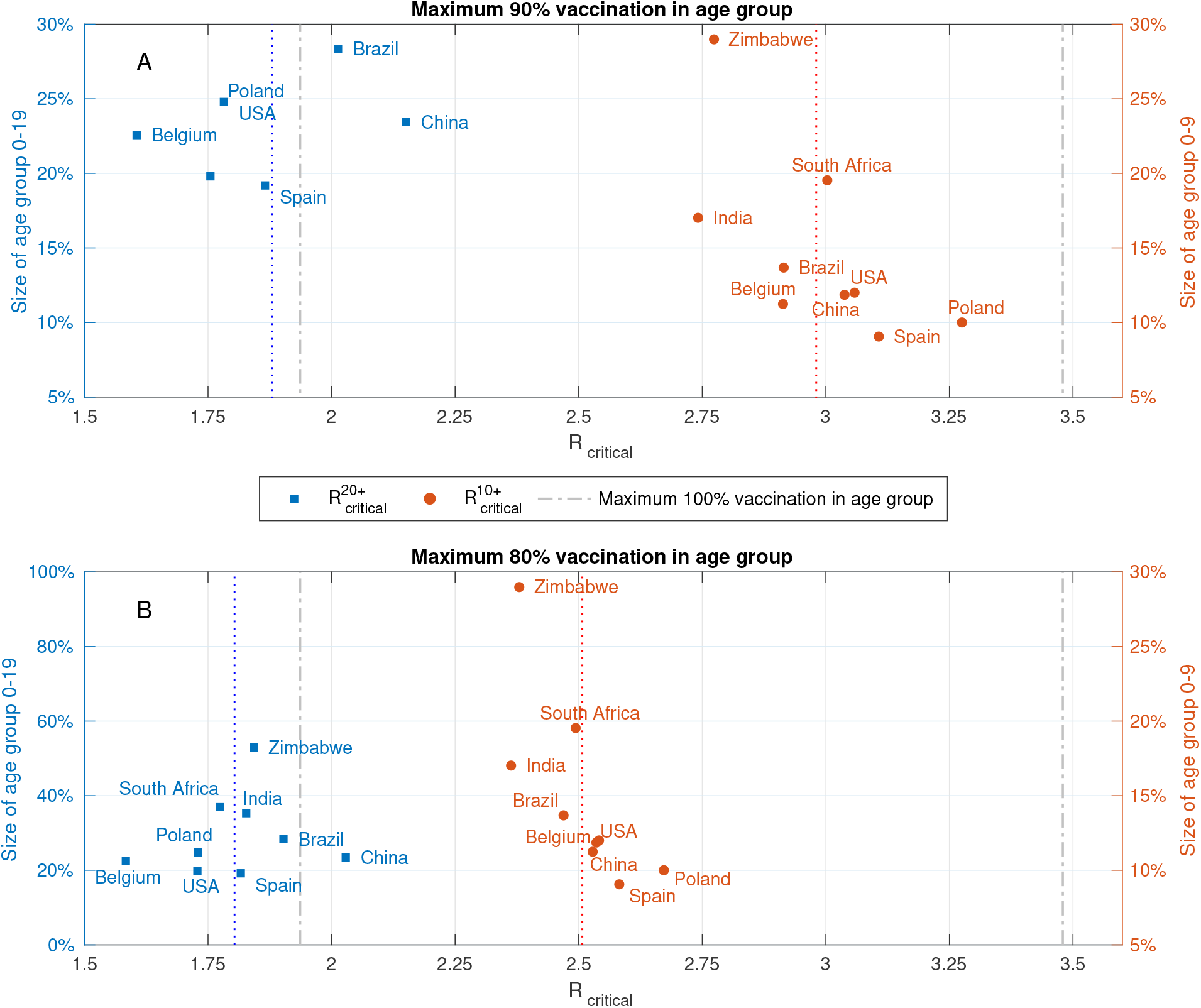
Effect of vaccine hesitancy on critical reproduction numbers. Reproduction numbers 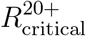 and 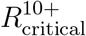 at which herd immunity cannot be achieved without vaccination of age groups 0 − 19 and 0 − 9, respectively. Computed for A: Maximum 90% vaccination in age group. B: Maximum 80% vaccination in age group. The gray curves in both plots correspond to the case when there is no constraint on the number of vaccines that can be allocated to an age group.

We observe that limiting vaccine coverage per age group to 90% results in an 220% increase in the minimal overall mortality that can be achieved and that further limiting vaccine coverage per age group to 80% results in an 330% increase in the minimal overall mortality that can be achieved, see Figure 17. These results strongly suggest that a key performance measure for the success of a vaccination campaign in reducing mortality should be vaccine coverage per age group, particularly in older age groups. In addition, we observe that the minimal number of infections that can achieved by proper allocation of vaccines is not as sensitive to the maximal possible vaccine coverage per age group. Indeed, we find that limiting vaccine coverage per age group to 90% results in 7% increase in the minimal overall mortality that can be achieved and that further limiting vaccine coverage per age group to 80% results in an 19% increase.

**Figure 17:**
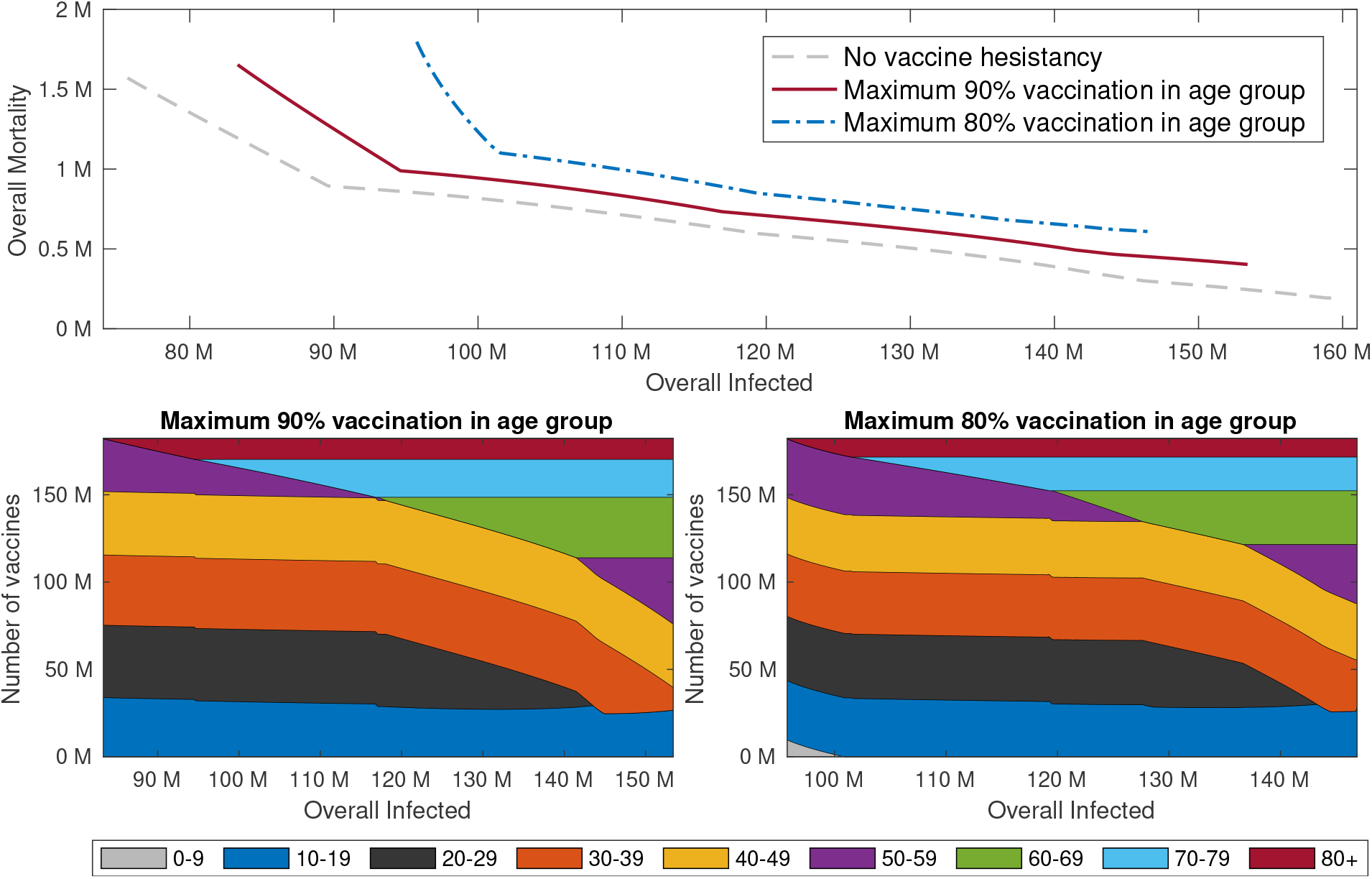
Pareto fronts for different vaccine hesitancy levels. Top graphs: Pareto fronts corresponding to a maximum of 90% (solid red), 100% (dashed gray) and 80% vaccine coverage per age group (dash-dotted blue). All other parameters are as in Figure 5. Bottom Graphs: Vaccine allocations along the corresponding Pareto fronts with 90% maximal vaccine coverage per age group (left) and 80% maximal vaccine coverage per age group (right).

### B.5 The effect of preexisting immunity in the population due to recovery

Most results presented in this work concerned population which is fully susceptible, unless vaccinated, at the end of the vaccination campaign. In practice, as of April 2021, the number of COVID-19 cases exceeded 130 million globally [34], many of which are recovered. We now study the effect of preexisting immunity in the population due to recovery. In particular, we consider cases in which 10% or 20% of the population are recovered and immune at the end of the vaccination campaign. We determine the distribution of the recovered cases among age groups in a way which is roughly equivalent to running a simulation without any vaccination until the recovered compartments reach the desired size. This is done by determining the distribution of the recovered cases according to the dominant eigenvector of the next generation matrix. A recovered individual is assumed to be fully immune to re-infection. Note, in comparison, that vaccinated individuals are assumed to be 80% immune to infection. In the computations below, we further assume that vaccines are not allocated to recovered cases.

We first consider the impact of preexisting immunity on vaccination coverage required for herd immunity. As expected, the leading order effect of 10% or 20% preexisting immunity, is that vaccine coverage *V*_threshold_ required to achieve herd immunity is reduced by 12% or 25%, respectively, see Figure 18. The differences between the reduction in *V*_threshold_ and the percent of population with preexisting immunity stem from the difference in the assumed immunity of recovered and vaccinated, and the fact that preexisting immunity is not optimally distributed.

**Figure 18:**
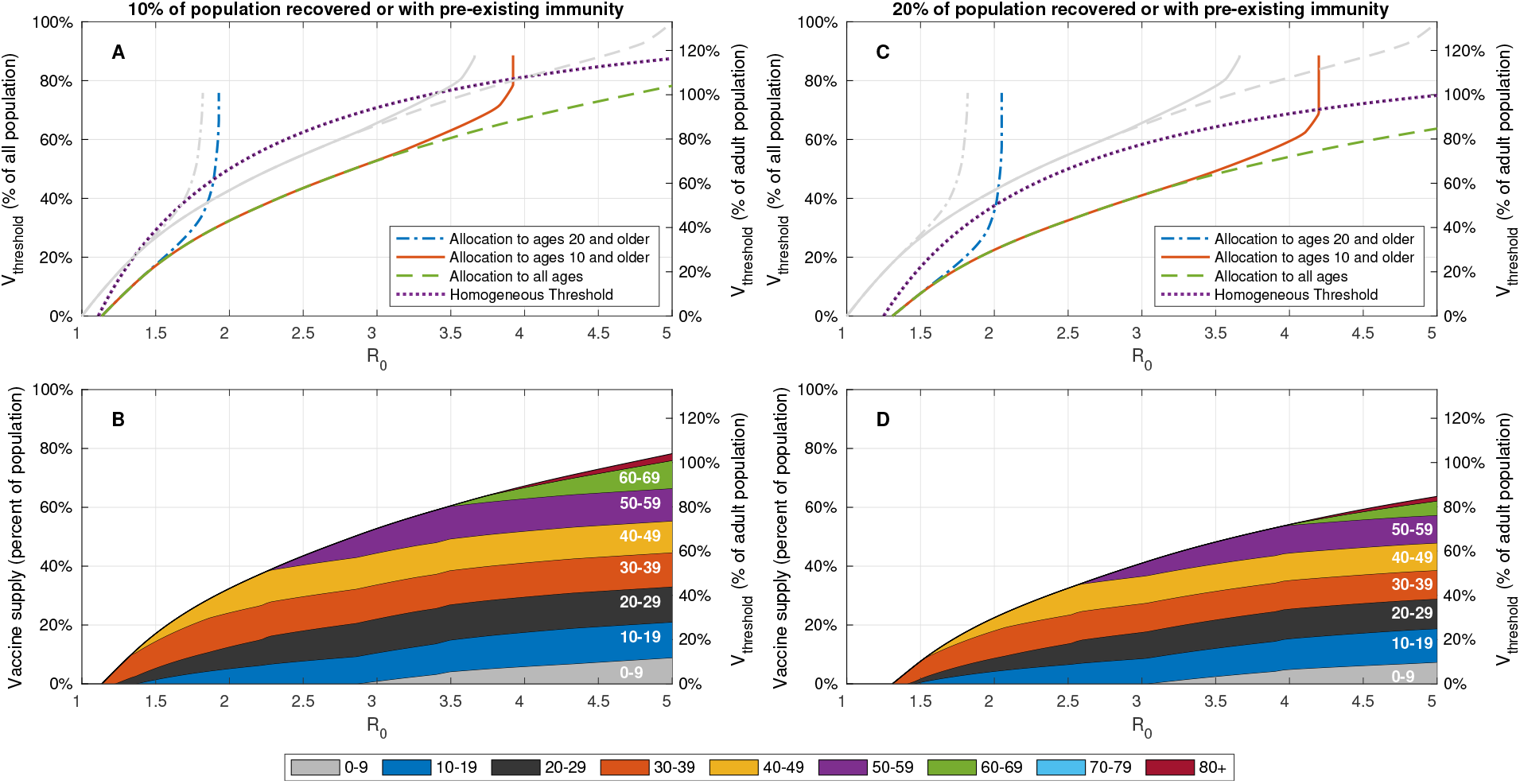
Impact of preexisting immunity on vaccination coverage required for herd immunity. A,C: Vaccine coverage *V*_threshold_ required to achieve herd immunity threshold as a function of the reproduction number *R*_0_ for the USA demography and contact structure. The gray curves corresponds to the case of no preexisting immunity. B: Vaccine allocations at which herd immunity is achieved at minimal vaccine coverage and when the entire population is eligible for vaccination.

The critical reproduction numbers at which herd immunity cannot be achieved without vaccination of age groups 0−9 or 0−19 increase with the percent of the individuals with preexisting immunity in these age groups. We compute the critical reproduction numbers for nine different countries, and observe that on average, prior immunization of 10% of the population results in the increase of roughly 7% in the critical reproduction numbers 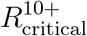 and 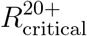. Similarly, prior immunization of 20% of the population results in an increase of about 15% in the critical reproduction numbers 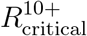 and 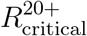 Next, we compute the Pareto front for the case in which 20% of the population is recovered or has prior immunity and vaccine coverage is 37% of the population (maximal coverage of 50% of population above the age of 20). Vaccines are allocated optimally taking into account the distribution of the recovered population. To study the effect of preexisting immunity, we also consider the case in which none of the population is recovered, but vaccine coverage is 60% of the population. The two scenarios are comparable in the sense that in both of them roughly half the population remains susceptible, where each vaccinated individual is counted and as 0.2 susceptible individual to account for 80% vaccine efficacy in blocking transmission. We observe that in the case of preexisting immunity the number of infections along the Pareto front increase by roughly 30% compared to a case of no preexisting immunity, while mortality is only marginally effected, see Figure 20. This significant difference stems from the fact that preexisting immunity is not optimally distributed. For example, the optimal allocation for minimizing infections gives rise to an overall number of 42 million infections (left end of dashed gray curve in Figure 20), while in the case of preexisting immunity the number of overall infections including the recovered population is bounded below by 66 million people (20% of the population) and reaches 95 million people.

**Figure 19:**
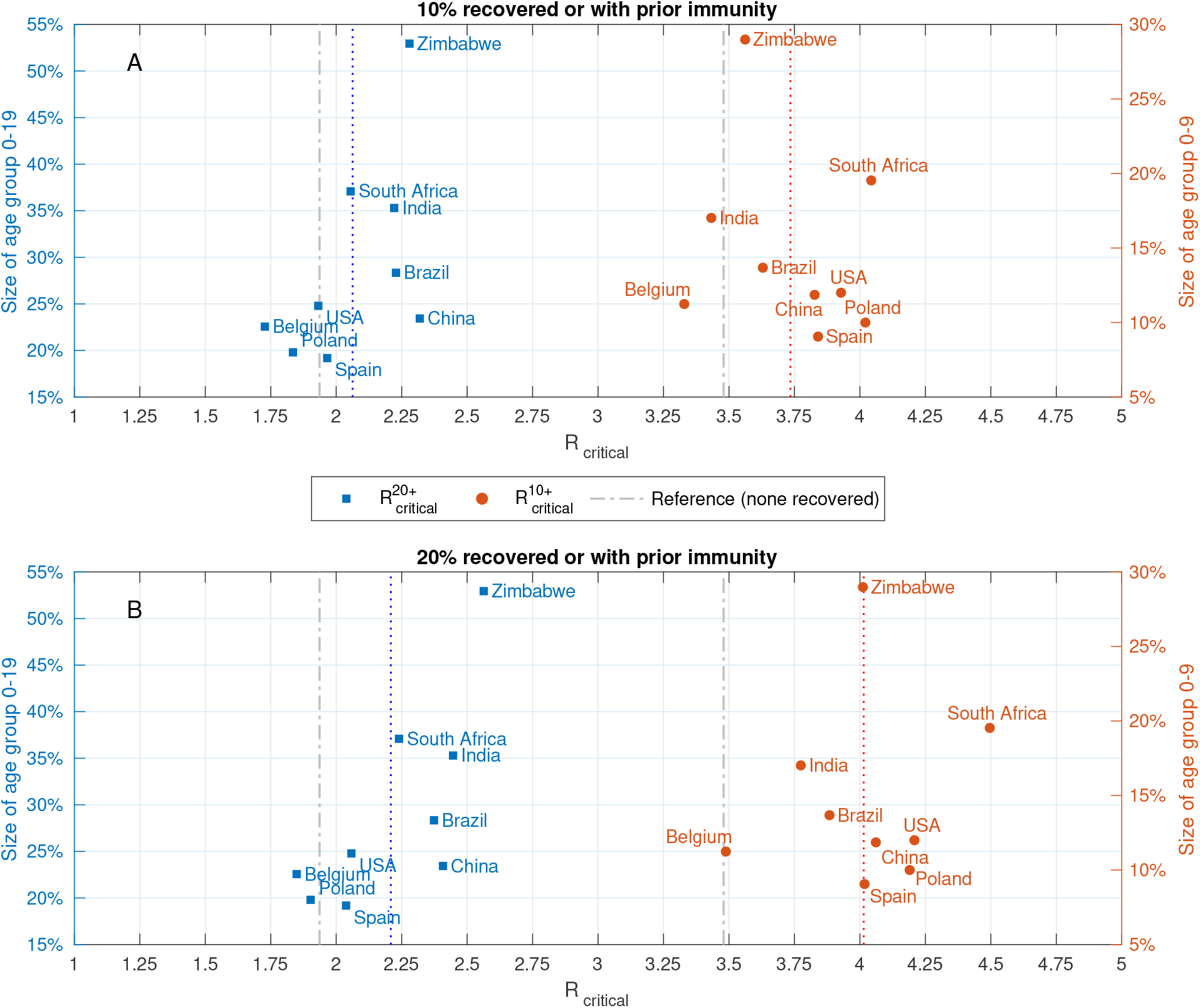
Impact on preexisting immunity on critical reproduction numbers. Reproduction numbers 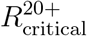 and 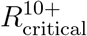 at which herd immunity cannot be achieved without vaccination of age groups 0 − 19 and 0 − 9, respectively. Computed for A: 10% of the population is recovered or has prior immunity. B: 20% of the population is recovered or has prior immunity. The gray curves in both plots correspond to the case when there is no prior immunity.

**Figure 20:**
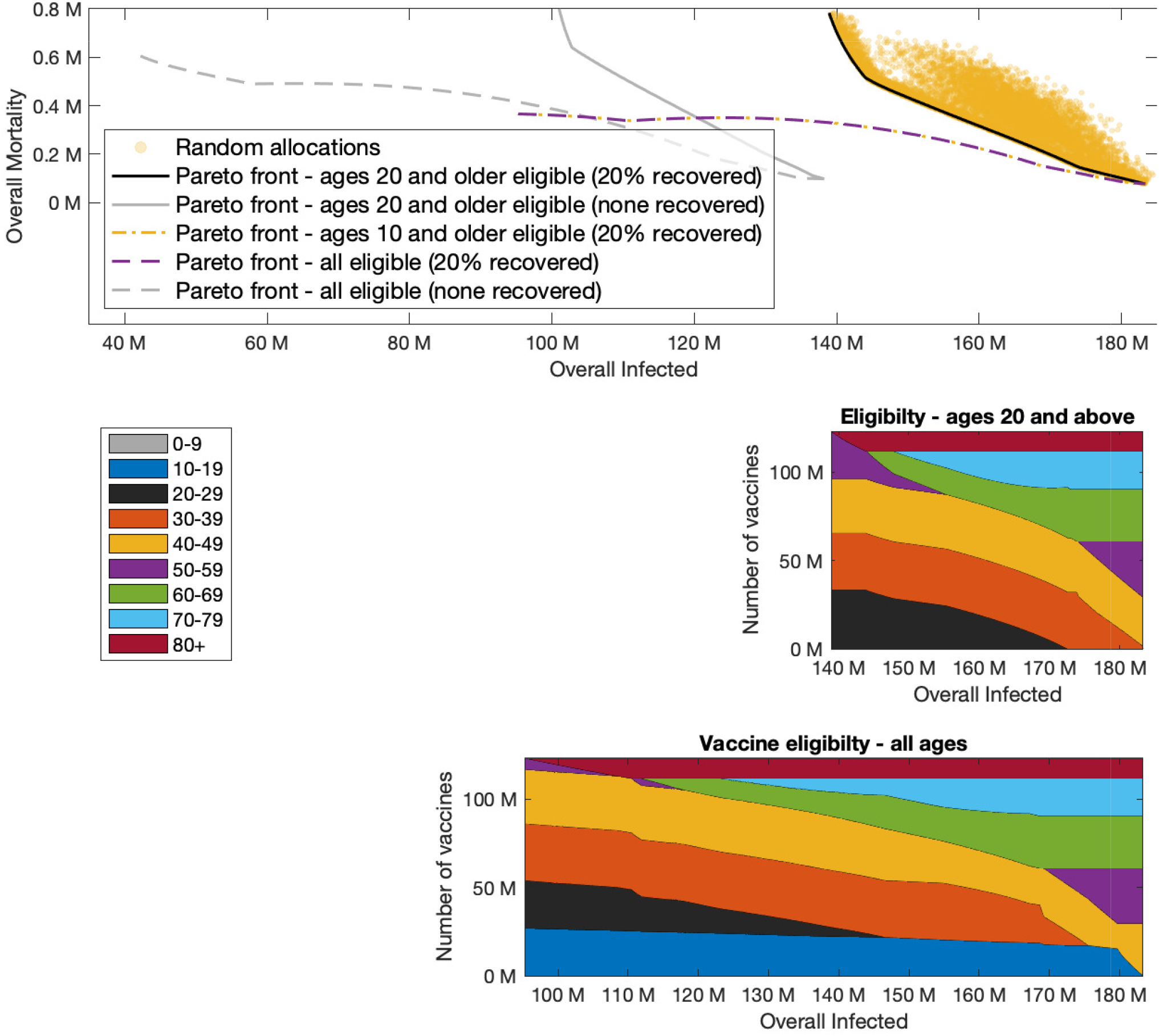
Pareto front with preexisting immunity. Pareto fronts computed for the case in which 20% of the population is recovered or has prior immunity and vaccine coverage is 37% of the population. The gray curves in the upper plot correspond to the case of no preexisting immunity and vaccine coverage of 60% of the population. Overall number of infected people includes the number of people recovered or with preexisting immunity.

## Notes

### Competing Interest Statement

The authors have declared no competing interest.

### Author Declarations

This work is based on the use of public aggregate data only and no medical intervention and is therefore exempt from institutional review board approval.

